# The changing face of nicotine use in England: age-specific annual trends, 2014-2024

**DOI:** 10.1101/2025.06.05.25329070

**Authors:** Sarah E. Jackson, Lion Shahab, Vera Buss, Harry Tattan-Birch, Sharon Cox, Eve Taylor, Jamie Brown

## Abstract

**Objective:** To examine age-specific trends in patterns of nicotine use in England between 2014 and 2024, including types of products used, exclusive and dual use of smoking and vaping, smoking frequency, and the smoking history of those who vape.

**Design:** Repeat monthly cross-sectional analysis of data from a nationally representative survey (the Smoking Toolkit Study).

**Setting:** England, 2014-2024.

**Participants:** 217,433 adults (≥18y).

**Main outcome measures:** Prevalence of (non-medicinal) nicotine use overall and by product type (combustible tobacco, e-cigarettes, heated tobacco products, and nicotine pouches), exclusive and dual use of smoking and vaping, daily versus non-daily smoking, and smoking history among those who vape. Estimates were stratified by age group (18-24, 25-34, 35-44, 45-54, 55-64, ≥65y).

**Results:** Nicotine use patterns varied markedly by age. Among 18-24-year-olds, vaping prevalence increased fivefold from 5.0% in 2014 to 25.0% in 2024 (prevalence ratio [PR]=5.00 [95%CI=4.18-5.91]), surpassing smoking by 2023. This contributed to an overall increase in nicotine use (26.1% to 36.5%; PR=1.40 [1.29-1.53]), despite declining smoking rates (25.3% to 19.9%; PR=0.79 [0.71-0.88]). In this age group, exclusive vaping became the most common mode of nicotine use, while nicotine pouch use also increased. Daily smoking declined substantially among 18-24-year-olds who smoked, with a shift toward non-daily smoking. Similar trends were observed among adults aged 25-44, though changes were smaller with increasing age. In older age groups (≥45), daily smoking declined modestly while vaping rose gradually, but there was little overall change in the prevalence of nicotine use. Most adults who vaped had a history of smoking, but the proportion who had never regularly smoked increased, particularly among 18-24-year-olds (4.3% to 34.3%; PR=7.98 [4.56-26.2]).

**Conclusions:** Generational shifts in nicotine use are occurring in England. Nicotine use has risen among young adults over the past decade, but they are increasingly moving away from daily cigarette smoking towards vaping or non-daily smoking. While older adults have also shown movement away from daily smoking, traditional smoking patterns remain more prevalent in this group. These trends suggest vaping may gradually replace smoking as the dominant form of nicotine consumption.

## Introduction

The landscape of nicotine use in England has changed substantially in recent years. For decades, nicotine consumption occurred almost exclusively through tobacco smoking (primarily cigarettes) and smoking prevalence had been declining steadily.^1^ The emergence and rapid uptake of electronic cigarettes (e-cigarettes or vapes)^2^ has since introduced a new dimension to population-level patterns of nicotine consumption. E-cigarettes offer a less harmful alternative to combustible tobacco products^3^ and initially, they were primarily used as a harm reduction tool by people who smoked.^2^ However, their growing daily use – particularly among young adults and individuals who have never smoked^2,4–6^ – has sparked debate about their longer-term public health impact. Debates have centred on the potential for vaping to support smoking cessation, while also potentially contributing to nicotine uptake in those who might not otherwise have used it.^7^ Other novel nicotine products have also been introduced to the market in recent years, including modern heated tobacco products (electronic devices that heat processed tobacco with the aim of avoiding combustion) and oral nicotine pouches (tobacco-free pouches placed in the mouth to release nicotine).^8^

The UK government’s Tobacco and Vapes Bill,^9^ which is currently progressing through parliament with cross-party support,^10^ aims to create a smokefree generation while addressing growing concerns around youth vaping. The bill proposes landmark measures, including a ban on all tobacco sales to anyone born on or after 1 January 2009, alongside powers to implement tighter restrictions on the marketing, packaging, and availability of vaping products and other tobacco-free nicotine products (e.g. pouches), subject to consultation and secondary legislation.^9^ As policymakers seek to strike a balance between supporting smoking cessation and preventing uptake of smoking and vaping, timely evidence on age-specific nicotine use trends is important to inform effective and proportionate regulation.

Understanding how patterns of smoking, vaping, and use of other nicotine products are evolving across age groups can provide insights into the future trajectory of nicotine use. Generational differences – shaped by historical exposures, social norms, and policy environments – reveal not only how behaviours have evolved, but also where they may be headed.^11–15^ In particular, the smoking and vaping patterns of today’s young adults are especially informative, offering a window into the future composition of the nicotine-using population.

This study therefore aimed to describe how nicotine use among adults in England has evolved over the past decade. Using nationally representative data from the monthly cross-sectional Smoking Toolkit Study, we examined age-specific annual trends in the prevalence and patterns of nicotine use between 2014 and 2024, including the types of products used and distinctions between exclusive and dual use of smoking and vaping, daily and non-daily smoking, and the smoking history of adults who vape.

## Methods

### Design

The Smoking Toolkit Study is an ongoing monthly cross-sectional survey designed to monitor trends in smoking in England.^16,17^ Full details of the study’s methodology are available elsewhere.^2,16,17^ Briefly, the study uses a hybrid of random probability and simple quota sampling to select a new sample of approximately 1,700 people aged 16 years and older across England each month. Raking is used to weight the sample to match the population in England.^16^ This profile is determined each month by combining data from the UK Census, the Office for National Statistics mid-year estimates, and the annual National Readership Survey.^16^ Comparisons with other national surveys and cigarette sales data have shown the survey produces nationally representative estimates of key sociodemographic and smoking indices.^16,18^

The survey began in November 2006 and has been conducted each month since, except for December 2008 and April 2020. Data were collected face-to-face up to the start of the Covid-19 pandemic and via telephone from April 2020 onwards; the two modes generally show good comparability.^19,20^ The lower age limit was raised to 18 years between April 2020 and December 2021.

The present analyses used data collected between January 2014 and December 2024 (the earliest and most recent complete years of data including vaping status available at the time of analysis). We restricted the sample to adults aged ≥18 years (the legal age of sale of tobacco and e-cigarettes) for consistency across the time series, and aggregated data annually for analysis.

### Measures

Smoking status was assessed by asking participants which of the following statements best applied to them: (a) I smoke cigarettes (including hand-rolled) every day; (b) I smoke cigarettes (including hand-rolled), but not every day; (c) I do not smoke cigarettes at all, but I do smoke tobacco of some kind (e.g. pipe, cigar or shisha); (d) I have stopped smoking completely in the last year; (e) I stopped smoking completely more than a year ago; (f) I have never been a smoker (i.e. smoked for a year or more). For analysis, responses were categorised as follows:

- Current smoking: (a)-(c)
- Daily cigarette smoking: (a)
- Non-daily cigarette smoking: (b)
- Exclusive non-cigarette tobacco smoking: (c)
- Recent former smoking: (d)
- Long-term former smoking: (e)
- Never regular smoking: (f)

Vaping status was assessed within several questions asking about use of a range of nicotine products. Participants reporting current smoking were asked ‘Do you regularly use any of the following in situations when you are not allowed to smoke?’ and those who reported cutting down ‘Which, if any, of the following are you currently using to help you cut down the amount you smoke?’; those reporting current smoking or recent former smoking were asked ‘Can I check, are you using any of the following either to help you stop smoking, to help you cut down or for any other reason at all?’; and those reporting long-term former smoking or never regular smoking were asked ‘Can I check, are you using any of the following?’. We defined current vaping as reporting using an ‘electronic cigarette’ or ‘Juul’ in response to any of these questions.

Use of heated tobacco products and nicotine pouches was assessed within the same questions as vaping status. We defined heated tobacco use as reporting using a ‘heat-not-burn cigarette (e.g. iQOS, heatsticks)’ and nicotine pouch use as reporting using ‘tobacco-free nicotine pouch/pod or white pouches that you place on your gum (e.g., Zyn, On!, Nordic Spirit, Velo, Lyft, Skruf)’ in response to any of these questions. As these products are relatively new, their use was not assessed across the entire period. Analyses of these variables was therefore limited to 2017-2024 for heated tobacco use and to 2021-2024 for pouch use (the only complete years of data to assess use of these products).

Nicotine use was defined as current smoking, vaping, heated tobacco use, or nicotine pouch use. We did not include the use of nicotine replacement therapy (NRT; e.g., nicotine patches, gum, lozenges, or inhalators) because these are regulated as medicinal rather than consumer products (available in England on prescription or over-the-counter).

Among those who smoked or vaped, exclusive smoking was defined as current smoking and no current vaping; exclusive vaping was defined as current vaping and no current smoking; and dual use was defined as both current smoking *and* current vaping.

Age was categorised as 18-24, 25-34, 35-44, 45-54, 55-64, and ≥65 years. Survey year was categorised as 2014 through 2024.

### Statistical analysis

Analyses were conducted using R v.4.4.1. The analyses were not pre-registered and should be considered exploratory. We excluded participants with missing data on age, smoking, or vaping. All analyses were run on weighted data, but sample sizes are reported unweighted.

Within each age group and year, we estimated: (i) the overall prevalence of nicotine use and the prevalence of smoking, vaping, heated tobacco use, and nicotine pouch use among adults; (ii) among adults who smoke or vape, the proportion exclusively smoking, exclusively vaping, or dual using; (iii) among adults who smoke, the proportion using each type and pattern of smoking (daily cigarette, non-daily cigarette, or exclusive non-cigarette tobacco); and (iv) among adults who vape, the proportion with each smoking status (current, recent former, long-term former, or never regular smoking). We presented results graphically, providing annual estimates with 95% confidence intervals (CI) within each age group in **Supplementary Files 1-4**. We also reported absolute estimates of the proportion of adults exclusively smoking, exclusively vaping, and dual using in **Supplementary File 2** and the proportion of adults smoking cigarettes daily, smoking cigarettes non-daily, and exclusively smoking non-cigarette tobacco in **Supplementary File 3**.

For each outcome assessed over the whole period (all except prevalence of heated tobacco and nicotine pouch use), we reported relative and absolute changes from the start to the end of the study period alongside 95% CIs calculated using bootstrapping (500 replications). Relative changes were reported as prevalence ratios (PR; calculated as prevalence in 2024 divided by prevalence in 2014; **Table 1**) and absolute changes as percentage point changes (calculated as prevalence in 2024 minus prevalence in 2014; **Table 2**).

**Table 1.**
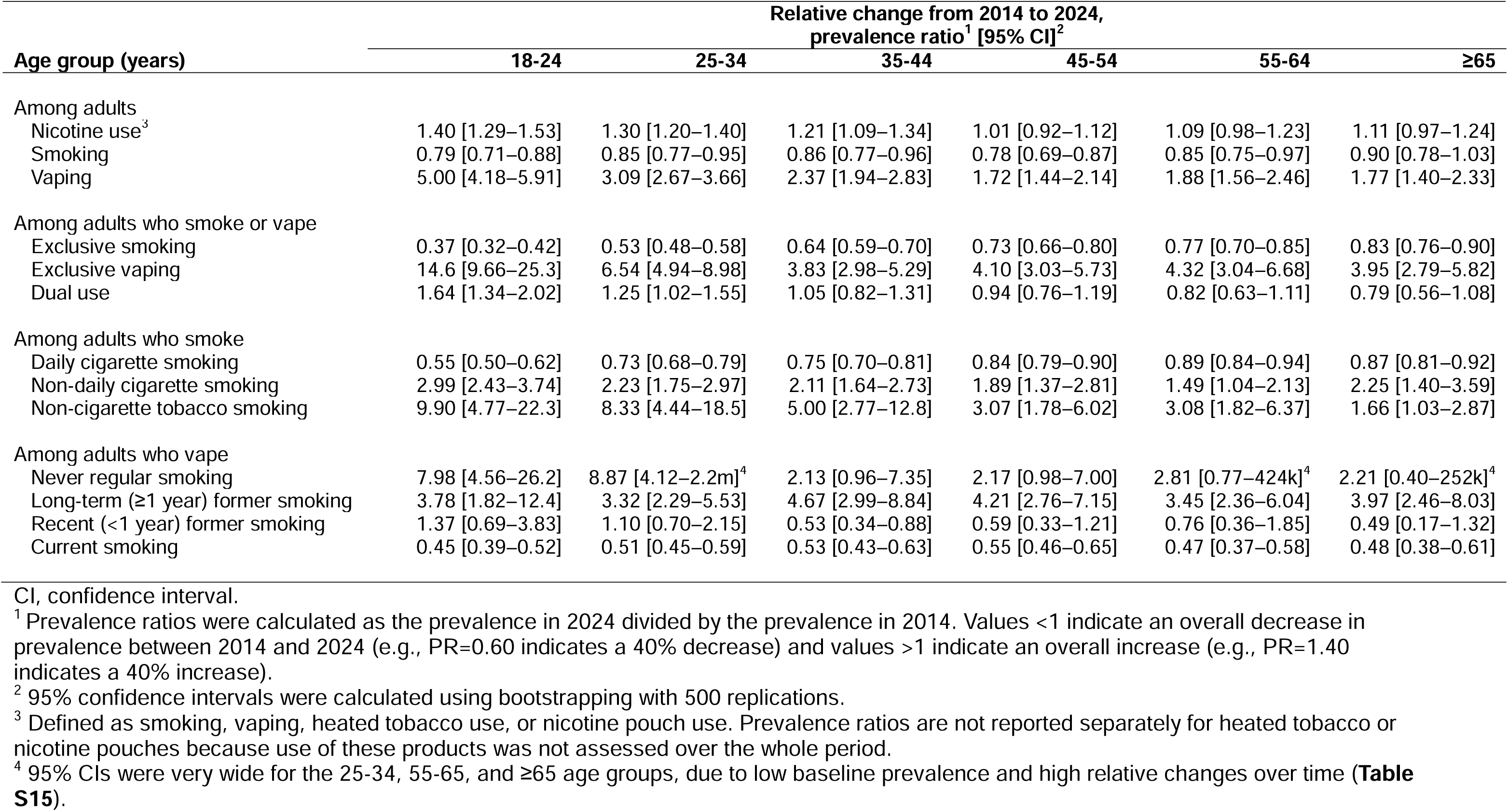
Age-specific relative changes in smoking and vaping patterns among adults in England, 2014 to 2024.

**Table 2.**
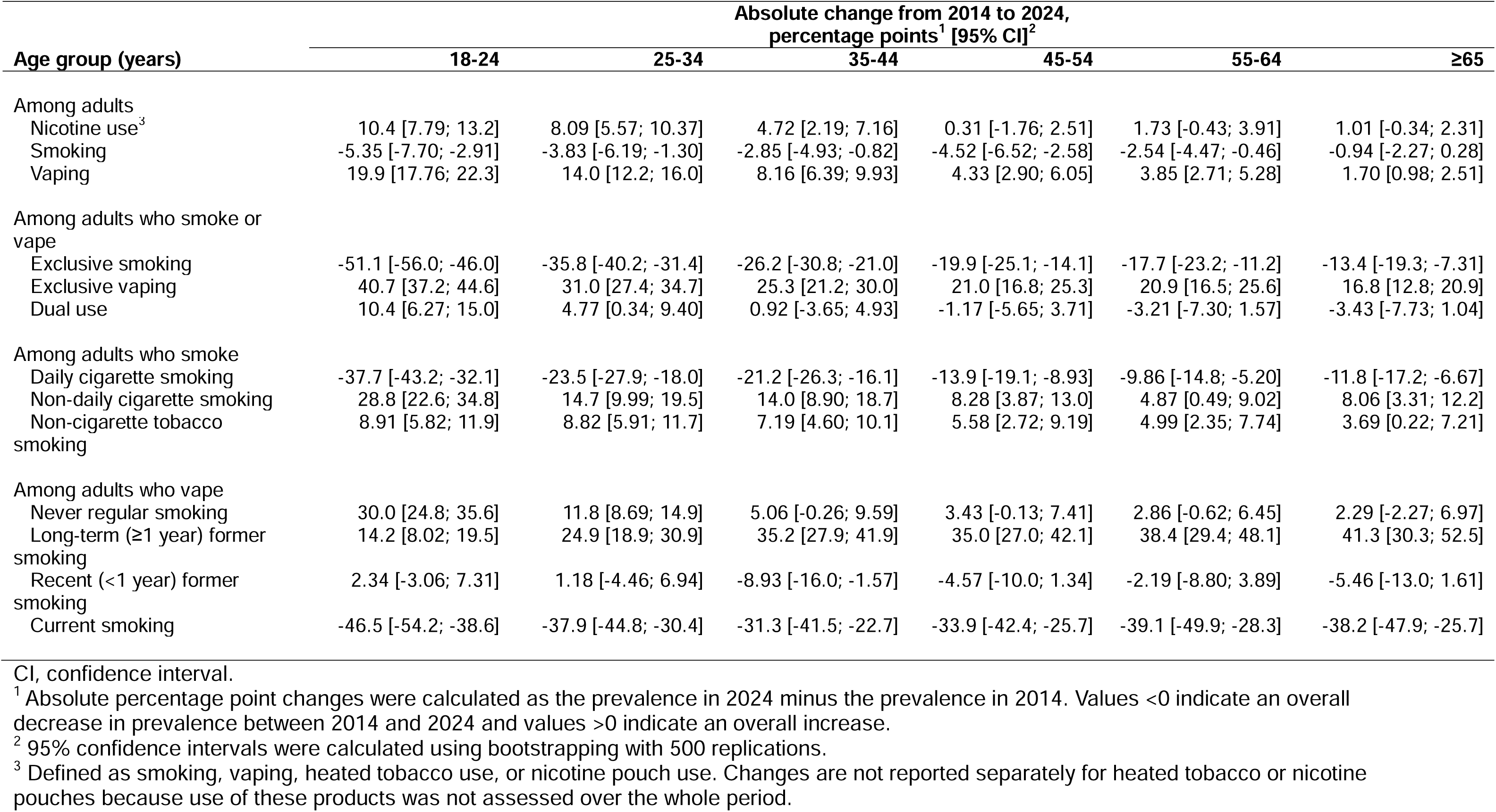
Age-specific absolute changes in smoking and vaping patterns among adults in England, 2014 to 2024.

## Results

There were 218,214 adults aged ≥18 years in England surveyed between January 2014 and December 2024. We excluded 784 (0.4%) with missing data on smoking or vaping status, leaving a final sample of 217,433 participants (weighted mean [SD] age = 49.6 [19.0] years; 49.8% women).

### Prevalence of nicotine use, smoking, vaping, heated tobacco use, and nicotine pouch use

Between 2014 and 2024, patterns of nicotine use varied considerably by age group (**Figure 1, Tables 1-2**). Among adults aged 18-24, overall nicotine use declined from 2014 to 2019, primarily due to a steady decrease in smoking, while vaping remained relatively stable. In 2020, smoking prevalence increased, before resuming its downward trend. Meanwhile, the decline in nicotine use reversed in 2021 as vaping surged. Vaping prevalence increased fivefold over the decade (from 5.0% in 2014 to 25.0% in 2024; PR=5.00 [95%CI 4.18–5.91]), overtaking smoking by 2023 and contributing to an overall rise in nicotine use (from 26.1% to 36.5%; PR=1.40 [1.29–1.53]) despite a marked decline in smoking (from 25.3% to 19.9%; PR=0.79 [0.71–0.88]). There was also a smaller increase in use of nicotine pouches (from 0.7% in 2021 to 3.4% in 2024), particularly since 2023, while use of heated tobacco products remained rare (<1%).

**Figure 1.**
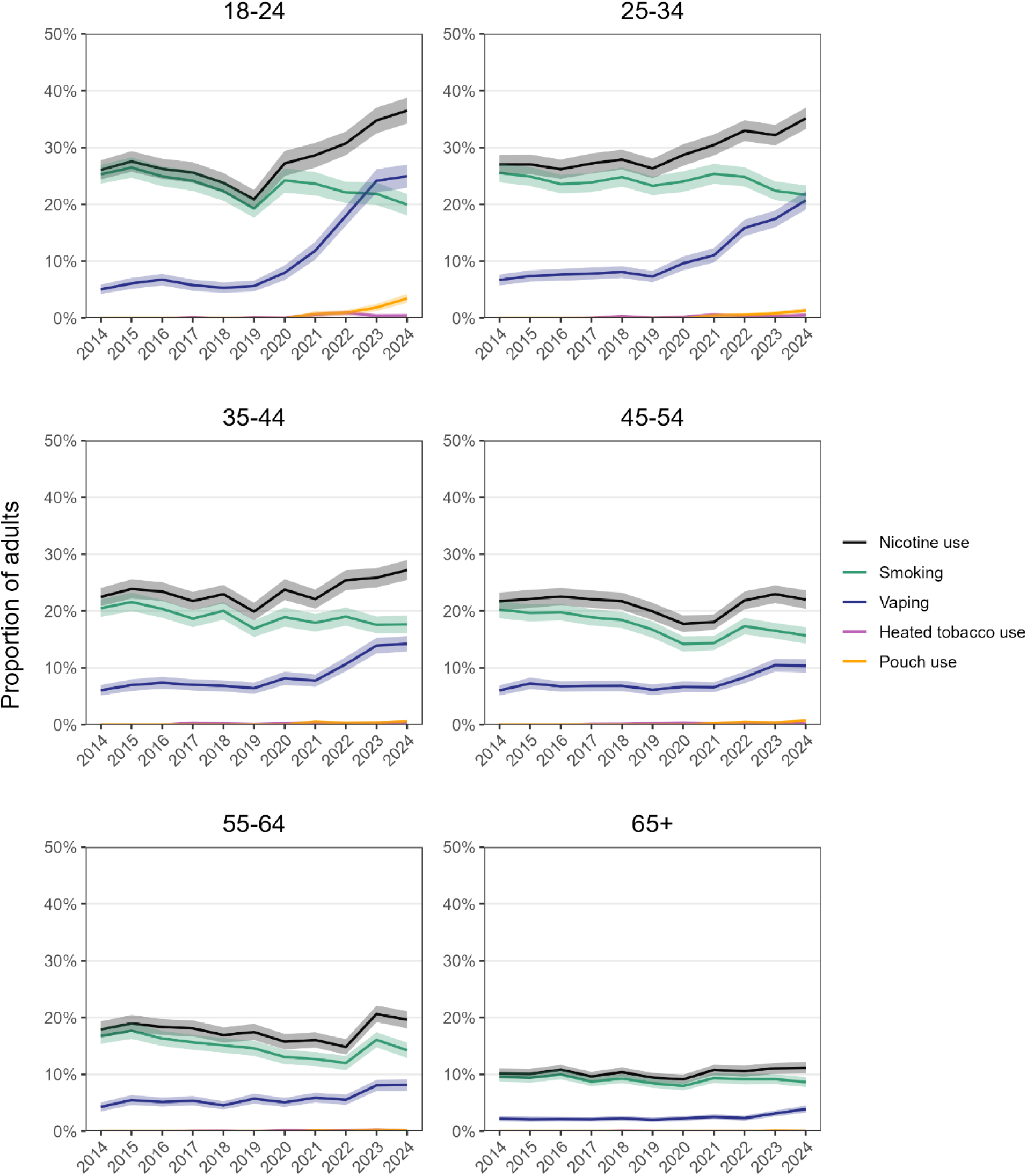
Prevalence of nicotine use, smoking, vaping, heated tobacco use, and nicotine pouch use among adults in England, 2014 to 2024. Lines represent weighted prevalence by year. Shaded bands represent 95% confidence intervals. Nicotine use is defined as smoking, vaping, heated tobacco use, or nicotine pouch use. Data are provided in tabular form in **Supplementary File 1**.

Similar, though less pronounced, changes were observed among adults aged 25-34. Smoking declined (from 25.5% in 2014 to 21.7% in 2024; PR=0.85 [0.77–0.95]), while vaping prevalence tripled (from 6.7% to 20.7%; PR=3.09 [2.67–3.66]), nearly reaching parity with smoking by 2024. The rise in vaping more than offset the decline in smoking, contributing to an overall rise in nicotine use (from 27.0% to 35.1%; PR=1.30 [1.20–1.40]). Use of heated tobacco products and nicotine pouches also increased, but remained rare (0.5% and 1.3%, respectively, in 2024).

Among those aged 35-44, comparable trends were observed, though changes were ever more modest. The overall prevalence of nicotine use rose only slightly (from 22.5% to 27.2%; PR=1.21 [1.09–1.34]). Smoking continued to decline (from 20.5% to 17.6%; PR=0.86 [0.77–0.96]), while vaping more than doubled (from 6.0% to 14.2%; PR=2.37 [1.94–2.83]), suggesting a slower but ongoing shift in product use. Use of heated tobacco products or nicotine pouches remained low and relatively consistent over time in this age group (0.4% and 0.6%, respectively, in 2024).

Among older age groups (≥45), nicotine use generally remained stable over the decade; more modest increases in vaping were offset by declining smoking rates, with minimal impact from heated tobacco products or nicotine pouches, which remained uncommon throughout. Among those aged 45-54, smoking declined (from 20.2% to 15.7%; PR=0.78 [0.69–0.87]), while vaping increased (from 6.0% to 10.3%; PR=1.72 [1.44–2.14]) but remained less prevalent than in younger groups. Among those aged 55-64, smoking followed a downward trajectory overall (from 16.8% to 14.2%; PR=0.85 [0.75–0.97]), though this was disrupted by a slight uptick in the final years of the study period (2022-2024), suggesting a potential reversal or plateau in progress. Meanwhile, vaping prevalence increased slowly (from 4.3% to 8.1%; PR=1.88 [1.56–2.46]) but remained less common than smoking. Among those aged ≥65, both smoking and vaping remained comparatively lower and more stable throughout the study period (_≤_10.0% and _≤_3.9%, respectively), although there was evidence of a gradual increase in vaping (from 2.2% to 3.9%; PR=1.77 [1.40–2.33]), predominantly in the latter part of the period.

### Patterns of exclusive and dual use of smoking and vaping

Disaggregating inhaled nicotine use into exclusive smoking, exclusive vaping, and dual use revealed further differences across age groups (**Figure 2, Tables 1-2**). Among 18-24-year-olds who smoked or vaped, the proportion who exclusively smoked fell sharply (from 80.7% in 2014 to 29.6% in 2024; PR=0.37 [0.32-0.42]), especially after 2020. Exclusive vaping increased in parallel (from 3.0% to 43.7%; PR=14.6 [9.66-25.3]), becoming the most common pattern of inhaled nicotine use in this age group. Dual use also increased but to a lesser extent (from 16.3% to 26.7%; PR=1.64 [1.34-2.02]). Among all 18-24-year-olds (i.e., not just those who smoked and vaped), the absolute prevalence of exclusive smoking fell from 21.0% to 10.5%, the prevalence of exclusive vaping increased from 0.8% to 15.5%, and the prevalence of dual use increased from 4.3% to 9.5% (**Supplementary File 2**).

**Figure 2.**
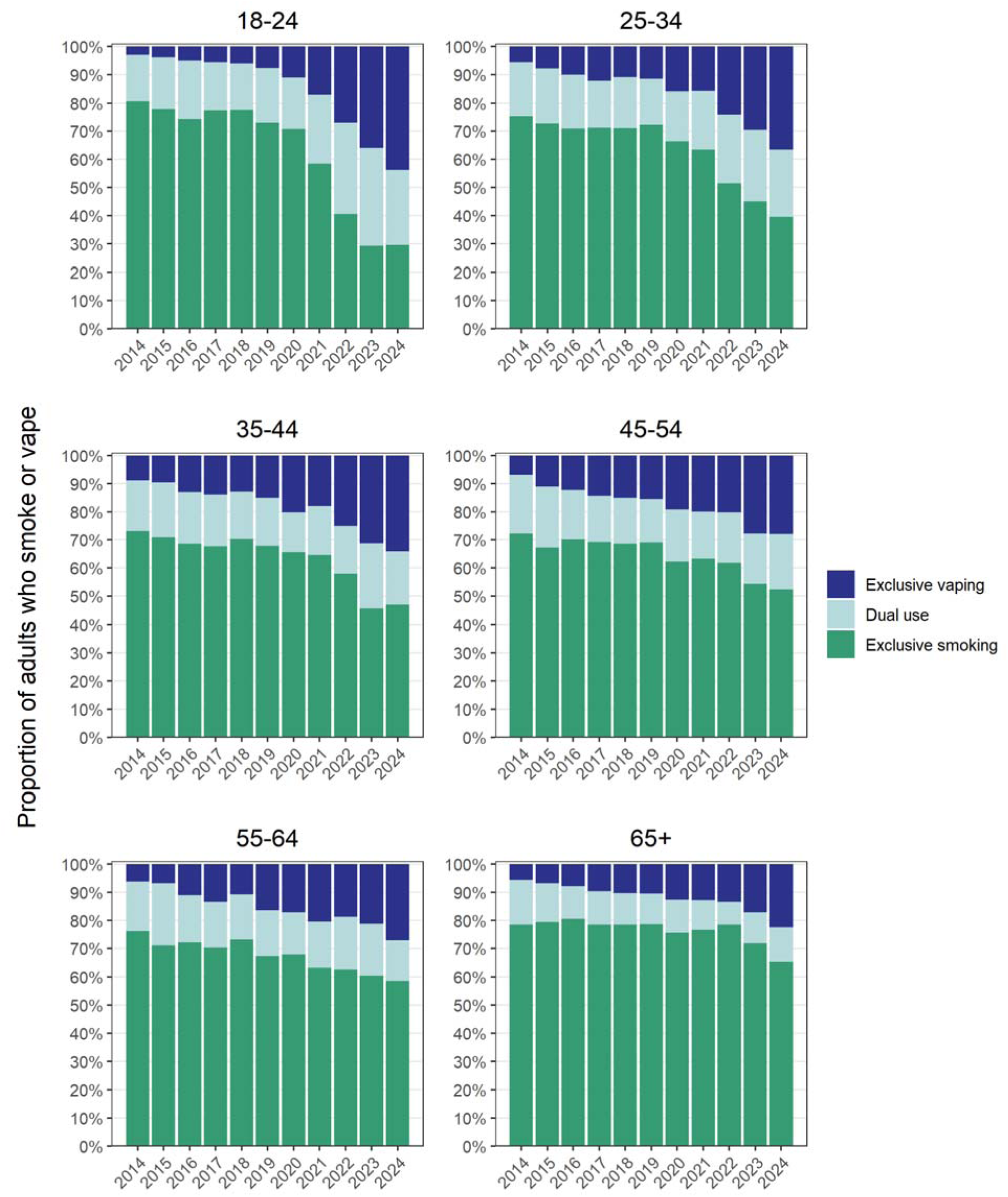
Prevalence of exclusive smoking, exclusive vaping, and dual use among adults who smoke or vape in England, 2014 to 2024. Estimates with 95% confidence intervals are provided in **Supplementary File 2,** along with corresponding figures and estimates for all adults (not just those who smoke or vape).

Adults aged 25-34 followed a similar trajectory, with substantial growth in exclusive vaping (from 5.6% to 36.6%; PR=6.54 [4.94-8.98]) alongside a decline in exclusive smoking (from 75.3% to 39.5%; PR=0.53 [0.48-0.58]). In the 35-44 and 45-54 age groups, exclusive smoking remained the predominant pattern of use throughout the period (46.9% and 52.4%, respectively, in 2024). However, exclusive vaping increased gradually (from 8.9% to 34.1%; PR=3.83 [2.98–5.29] and from 6.8% to 27.9%; PR=4.10 [3.03–5.73], respectively), especially from 2021 onward. In the older groups (55-64 and ≥65), exclusive smoking remained dominant (58.5% and 65.2%, respectively, in 2024), but there was still a notable increase in exclusive vaping over time (e.g., from 5.7% in 2014 to 22.5% in 2024 among those aged ≥65; PR=3.95 [2.79-5.82]), with relatively limited dual use in the oldest group (≥65) compared with other age groups.

### Smoking type and pattern among adults who smoke

As the prevalence of smoking declined, there were also shifts in the type and pattern of tobacco use among adults who smoked (**Figure 3, Tables 1-2**). In 2014, across all ages, the majority who smoked reported smoking cigarettes daily (range: 84.5-88.1%). However, among 18-24-year-olds who smoked, the proportion smoking cigarettes daily declined sharply (from 84.5% in 2014 to 46.7% in 2024; PR=0.55 [0.50-0.62]), particularly after 2020, while non-daily cigarette smoking increased (from 14.5% to 43.3%; PR=2.99 [2.43-3.74]). By 2023, fewer than half (44.8%) of 18-24-year-olds who smoked reported daily cigarette smoking. The proportion who did not smoke cigarettes at all but smoked other forms of tobacco increased substantially between 2019 and 2023 (from 3.5% to 14.4%), then declined slightly (to 9.9%) in 2024. Among all 18-24-year-olds (i.e., not just those who smoked), the absolute prevalence of daily cigarette smoking decreased from 21.4% to 9.3% between 2014 and 2024, while non-daily cigarette smoking increased from 3.7% to 8.6% and exclusive non-cigarette tobacco smoking increased from 0.3% to 2.0% (**Supplementary File 3**).

**Figure 3.**
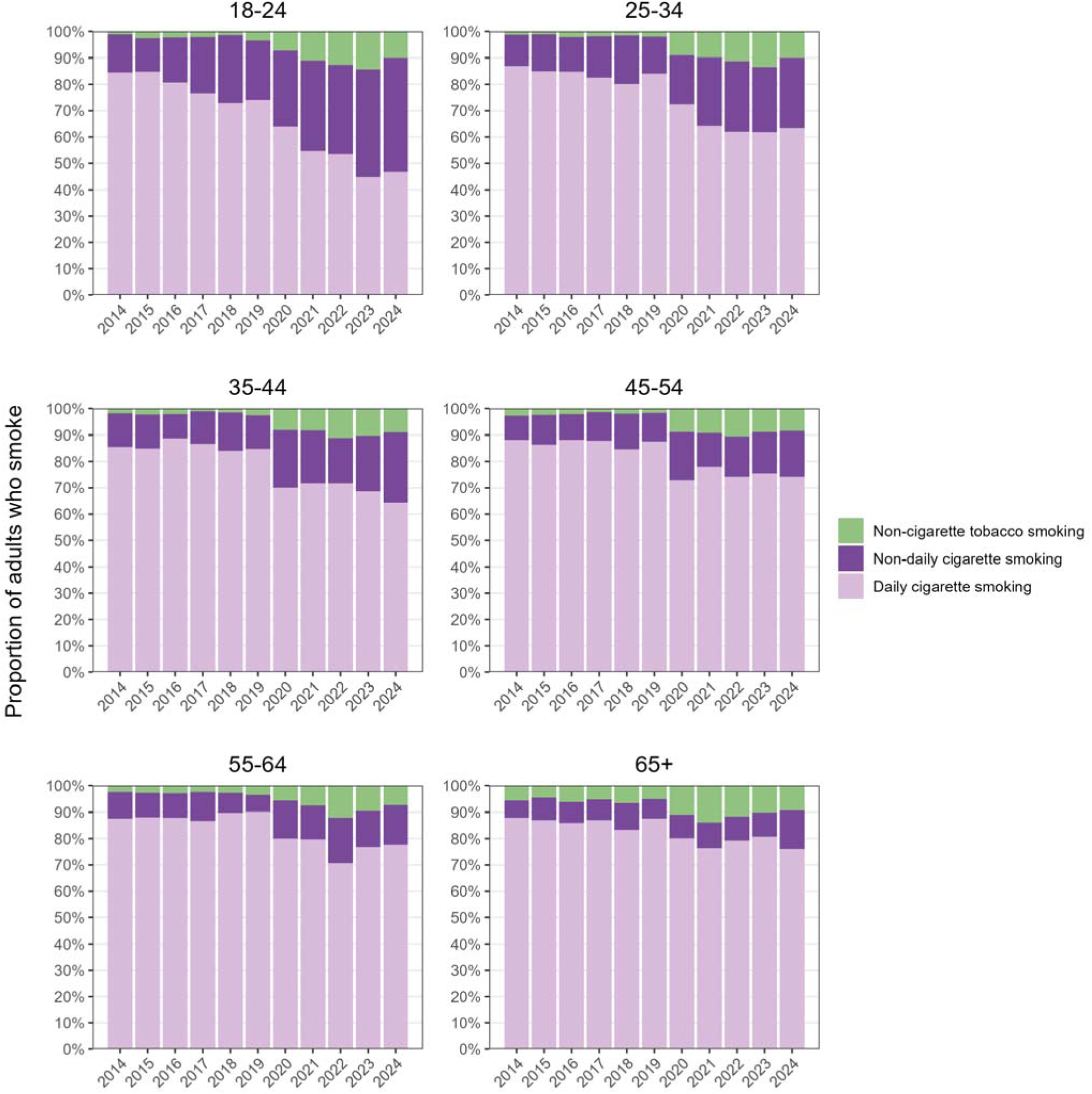
Smoking type and pattern among adults who smoke in England, 2014 to 2024. Estimates with 95% confidence intervals are provided in **Supplementary File 3,** along with corresponding figures and estimates for all adults (not just those who smoke).

Patterns were similar among adults aged 25-34 and 35-44, with a gradual decline in daily smoking accompanied by increases in both non-daily cigarette smoking and non-cigarette tobacco smoking. However, these changes were less pronounced than among the youngest group. In 2024, around two-thirds of those who smoked reported smoking cigarettes daily (63.4% and 64.4% among those aged 25-34 and 35-44, respectively).

In the older (45-54, 55-64, and ≥65) age groups, daily cigarette smoking remained the predominant pattern throughout the study period, with only modest increases in non-cigarette tobacco smoking and especially non-daily smoking. Compared with younger adults, these groups showed more stability in smoking behaviour, with slower transitions away from daily cigarette use. In 2024, around three-quarters who smoked still smoked cigarettes daily (74.2%, 77.7%, and 76.1% among those aged 45-54, 55-64, and ≥65, respectively). Declines in the absolute prevalence of daily smoking between 2014 and 2024 were ever more modest with increasing age (**Supplementary File 3**).

### Smoking history among adults who vape

As the prevalence of vaping increased, there were changes in the composition of the vaping population in terms of their smoking history (**Figure 4, Tables 1-2**). Across all age groups and years, the majority of adults who vaped had a history of smoking. However, the proportion of those who vaped who reported current smoking declined steadily from 2014 to 2024 in all age groups.

**Figure 4.**
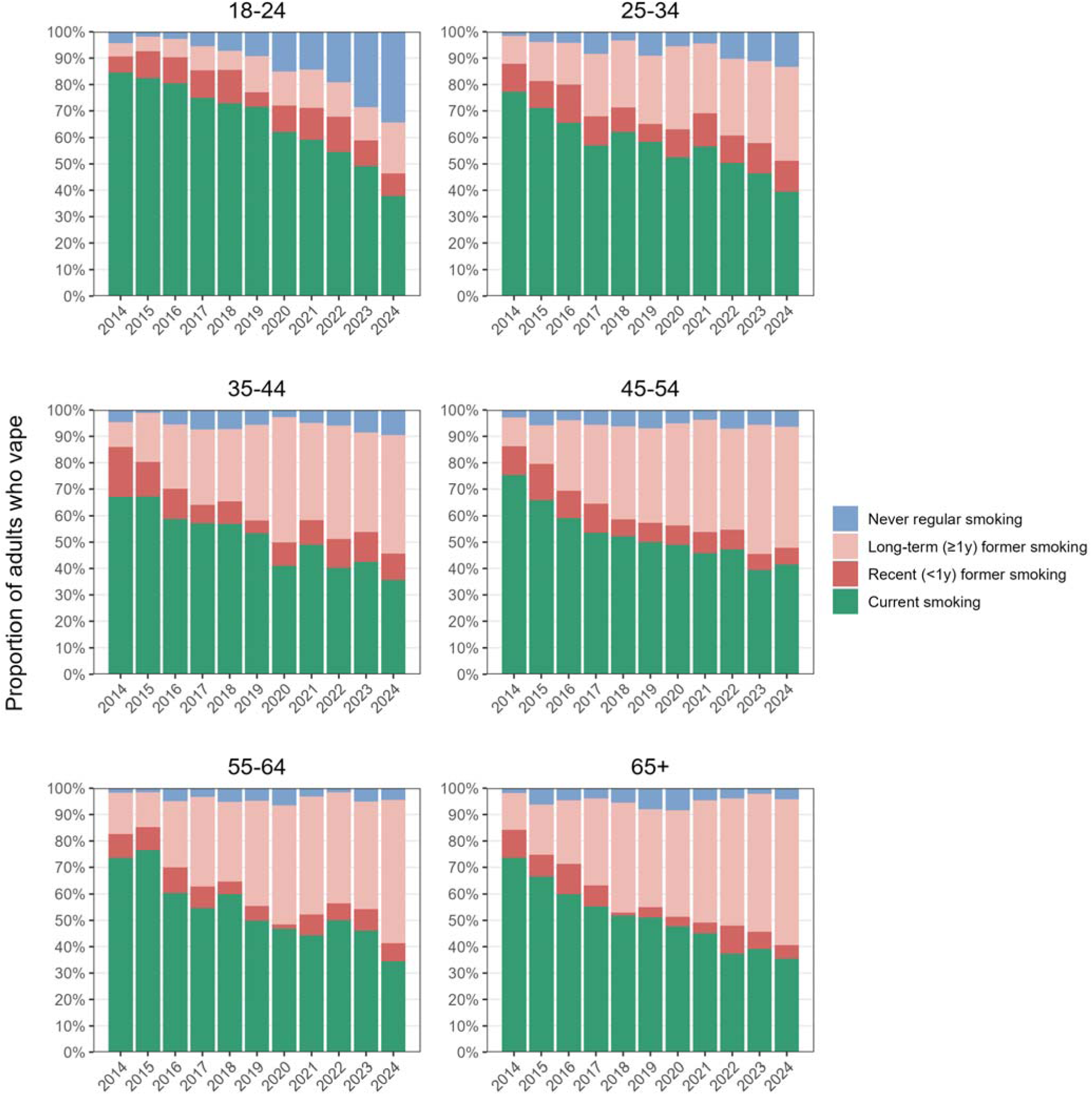
Smoking history among adults who vape in England, 2014 to 2024. Estimates with 95% confidence intervals are provided in **Supplementary File 4**.

Among 18-24-year-olds who vaped, this was largely driven by an increase in the proportion who had never regularly smoked (from 4.3% in 2014 to 34.3% in 2024; PR=7.98 [4.56–26.2]; +30.0 [24.8–35.6] percentage points), with this subgroup growing to a similar size as those who reported current smoking by 2024 (37.9%). There was also an increase, albeit smaller, in the proportion who reported long-term former smoking (i.e., quit ≥1 year ago; from 5.1% to 19.3%; PR=3.78 [1.82– 12.4]; +14.2 [8.02–19.5] percentage points).

Among those aged 25-34 who vaped, a similar pattern was observed, but with a smaller absolute increase in the proportion who had never smoked (from 1.5% to 13.3%; +11.8 [8.69–14.9] percentage points) and a larger absolute increase in the proportion who reported long-term former smoking (from 10.7% to 35.5%; +24.9 [18.9–30.9] percentage points).

In the older age groups (35-44, 45-54, 55-64, and ≥65) who vaped, the decline in the proportion who currently smoked was mostly offset by increases in the proportion who reported long-term former smoking. However, there were still increases in the proportion who had never regularly smoked, even in the oldest age group (≥65; from 1.9% to 4.2%), although the 95% CIs around this change indicate some uncertainty (+2.29 [-2.27; 6.97] percentage points).

## Discussion

This study presents a comprehensive picture of changing patterns of nicotine use among adults of different ages in England between 2014 and 2024, revealing substantial generational shifts.

The most striking changes occurred among younger adults (<45 years) where daily smoking declined markedly, particularly among 18-24-year-olds. Meanwhile, vaping increased sharply and surpassed smoking by 2023 in 18-24-year-olds. This coincided with rising overall nicotine use, driven by growth in exclusive vaping and, to a lesser extent, dual use and nicotine pouch uptake. Use of heated tobacco products remained rare. Similar trends were observed among adults aged 25-44, though changes were smaller with increasing age. These shifts likely reflect the growing appeal and availability of novel nicotine products, in particular e-cigarettes and nicotine pouches, which have been marketed in ways that particularly appeal to younger consumers; for example, endorsement by social media influencers, highlighting a range of appealing flavours, modern product designs, and lifestyle appeal.^22,23^ Although younger adults did not initially show higher uptake of vaping than older age groups, recent trends suggest that these products have become increasingly embedded in youth culture.^22,24^

Changes in nicotine use were less pronounced among adults aged 45-54 years. While vaping prevalence increased and smoking declined, these shifts did not lead to large changes in overall nicotine use. Exclusive smoking remained the dominant pattern of use throughout, and the majority of those who smoked in these age groups continued to report daily cigarette use. Adults aged ≥55 exhibited even more stability in nicotine use patterns, with smaller increases in vaping and more modest declines in smoking. This relative stability may reflect longer smoking histories, higher levels of nicotine dependence, and greater resistance to behaviour change (including less interest in trying new products) among older adults.^15,25,26^ The way vapes are marketed and portrayed may also mean they are less appealing to older age groups.^27^ Alternatively, self-selection pressures (such as healthy survivor effects), resulting in a more homogenous group less motivated to quit smoking, may contribute to the relative lack of change in this age group. Nevertheless, even in the oldest age group, vaping prevalence increased, and exclusive vaping became more common, indicating gradual but ongoing changes in older age groups.

Across all age groups, daily smoking declined more sharply than smoking overall. This pattern was particularly pronounced among young adults (18-24), among whom daily smoking prevalence fell to below 10% and non-daily smoking became the predominant pattern among those who smoked.

This indicates a meaningful reduction in the most harmful form of nicotine consumption.^3^ These findings suggest that vaping has not only displaced smoking for many young adults but may also have contributed to changes in the pattern and intensity of tobacco use among those who continue to smoke. Recent data (also from the Smoking Toolkit Study) show there has been a shift away from more regular smoking toward more regular vaping since 2016 among adults who both smoke and vape, and that daily vaping with non-daily smoking is a more common pattern of dual use among those who are younger.^28^ These shifts may also potentially reflect changes in social norms, whereby smoking – particularly daily smoking – has become less visible and less socially acceptable in younger cohorts, while vaping has gained traction as the normative form of nicotine use.^29^

Another notable finding was the evolving smoking history profile of adults who vape. Across all age groups, the proportion who reported former smoking increased, indicating they are being used as a tool for successful smoking cessation. Many people continue vaping for prolonged periods after using e-cigarettes to stop smoking,^30,31^ so as people have steadily quit smoking with e-cigarettes this group has grown. While most vapers continued to have a history of smoking, there were also increases in the proportion who had never regularly smoked, particularly in the youngest age group. By 2024, more than one in three 18-24-year-olds who vaped had never regularly smoked. This suggests that a growing number of young adults are initiating nicotine use with vaping, rather than transitioning from smoking. This shift likely reflects both displacement of smoking initiation in younger generations and the growing appeal of vaping as a standalone behaviour, culturally distinct from smoking. It is probable that some people who vape but have never regularly smoked may have otherwise initiated regular smoking in the absence of vaping,^6^ which would confer greater health risks.^3^ However, with the overall prevalence of nicotine use increasing to levels not seen since the 1990s,^32^ there will also be some young adults taking up vaping who would not have otherwise smoked.^5,6^ This will expose them to more risk than if they had not used any nicotine products at all.^3^ The emergence of newer products, such as e-cigarettes and nicotine pouches, may contribute to this trend, as they offer a less harmful alternative to traditional smoking,^3,25^ and are more socially acceptable^33^, cheaper, and more discreet.^34,35^ These features may make them attractive not only to people who smoke and seek alternatives, but also to individuals who may never have otherwise used nicotine. While these products may help reduce smoking rates, they also create new entry points for nicotine use, potentially broadening the user base and driving up overall prevalence even as smoking declines. The presence of people who vape but have never regularly smoked in older age groups raises important questions about how and why people are entering the nicotine market later in life. These individuals may provide a clearer view of new recruitment into nicotine use, given they were teens and young adults (the periods of life when smoking initiation is most common^36^) in the pre-vaping era.

The policy implications of these trends in nicotine use will depend on the ultimate goal of regulation. Traditionally, tobacco control efforts have focused on reducing tobacco-related harm, with an emphasis on lowering smoking prevalence and its associated health risks.^37,38^ However, the recent rapid growth in e-cigarette use and the emergence of other alternative nicotine products have led to the suggestion of a broader objective: a nicotine-free society.^39,40^ Our findings suggest such a goal requires nuanced consideration. While vaping appears to be playing a role in accelerating declines in daily cigarette smoking – especially among young adults – it is also contributing to a rise in overall nicotine use. This dual effect underscores the complexity of the evolving nicotine landscape and suggests policies must strike a balance between minimising harm and preventing new uptake. Given e-cigarettes expose users to much lower numbers and levels of toxicants compared to combustible tobacco,^3^ a continued shift in nicotine use from smoking toward vaping and other less harmful products would likely yield a net public health benefit^41^ – even if the total number of nicotine users increases. By reducing exposure to the most dangerous forms of nicotine consumption,^3^ such a shift could lead to meaningful reductions in smoking-related disease and mortality at the population level. In this case, prioritising harm reduction over reducing uptake of nicotine may be the most effective strategy for safeguarding public health. However, if these novel products confer a risk reduction that is counterbalanced by an increase in prevalence of use, then there could be population harms.

Current patterns of nicotine use in England present a unique opportunity in the history of tobacco control, with a large number of adults who smoke non-daily already using alternative nicotine products. This group could be well-positioned to quit smoking entirely, but widespread misperceptions about the relative harms of smoking and alternatives^42^ may hinder smoking cessation efforts. Public health messaging that clearly communicates the relative harms of smoking and alternative nicotine products is crucial for enabling people to make informed decisions about the products they use.

This study has several strengths. It uses large-scale, nationally representative data collected consistently over a decade, capturing a period of substantial change in nicotine use. Disaggregation by product type and smoking history provides a more detailed understanding of user profiles. However, there were also limitations. While the observed progression of nicotine use patterns across age groups is suggestive of a cohort effect, our analysis cannot definitively separate age, period, and cohort influences. Formal age-period-cohort modelling was not feasible at this stage because many of the trends we documented have unfolded within a short time frame but may be a useful avenue for future studies over the longer-term. Despite the large sample, 95% CIs were wide for estimates of rarer outcomes. The cross-sectional design means causal relationships cannot be established. For example, while increases in vaping coincided with declines in smoking, the extent to which vaping directly contributed to these changes or whether other factors (e.g., tobacco control policies, shifts in social norms) played a more significant role remains unclear. In addition, the repeat cross-sectional design meant we could not assess within-person transitions over time, such as whether dual users ultimately quit smoking. Finally, we did not explore the underlying motivations or reasons for changes in nicotine use behaviours, such as the influence of peer groups, marketing, or public health policies. Understanding these drivers would offer valuable insights for policymakers looking to design interventions that are responsive to changing patterns of nicotine use.

Together, these patterns suggest a rapidly changing nicotine landscape in England. Vaping is increasingly decoupled from smoking, particularly among younger and emerging middle-aged adults, and may continue to rise in prevalence among those who have never regularly smoked tobacco as current cohorts age. If current trends persist, and absent significant policy change, the population-level burden of daily cigarette smoking may continue to decline as younger cohorts – already exhibiting low daily cigarette smoking rates – age into older adulthood. The delayed but visible rise in vaping among those who have never regularly smoked across successive age bands hints at a future nicotine market that is less combustible, but potentially broader in terms of user base than it has been in the past decade. This evolving landscape may be influenced by ongoing legislative efforts, such as the Tobacco and Vapes Bill. Understanding these trends is critical for informing future policy, enabling it to respond effectively to the evolving dynamics of nicotine consumption.

## Data Availability

Data are available on request.

## Declarations

### Ethics approval

Ethical approval for the STS was granted originally by the UCL Ethics Committee (ID 0498/001). Participants provide informed consent to take part in the study, and all methods are carried out in accordance with relevant regulations. The data are not collected by UCL and are anonymised when received by UCL.

