## Supplementary file 1 for "The changing face of nicotine use in England: age-specific annual trends, 2014-2024"

### Table S1. Nicotine use prevalence by age group and year

|  | **Nicotine use, % [95% confidence interval]** | | | | | | | | | | |
| --- | --- | --- | --- | --- | --- | --- | --- | --- | --- | --- | --- |
|  | **2014** | **2015** | **2016** | **2017** | **2018** | **2019** | **2020** | **2021** | **2022** | **2023** | **2024** |
| 18-24 | 26.1  [24.4–27.8] | 27.5  [25.7–29.4] | 26.2  [24.5–28.0] | 25.6  [23.8–27.4] | 23.8  [22.0–25.5] | 20.9  [19.2–22.5] | 27.2  [25.0–29.4] | 28.6  [26.5–30.8] | 30.7  [28.7–32.8] | 34.8  [32.5–37.0] | 36.5  [34.2–38.8] |
| 25-34 | 27.0  [25.4–28.7] | 27.0  [25.3–28.8] | 26.2  [24.5–27.9] | 27.2  [25.4–29.0] | 27.9  [26.1–29.6] | 26.3  [24.6–28.0] | 28.7  [26.8–30.6] | 30.5  [28.6–32.3] | 33.0  [31.1–34.8] | 32.2  [30.4–34.0] | 35.1  [33.3–37.0] |
| 35-44 | 22.5  [20.9–24.1] | 23.8  [22.2–25.5] | 23.4  [21.7–25.0] | 21.7  [20.1–23.3] | 22.9  [21.3–24.6] | 19.9  [18.3–21.4] | 23.7  [21.9–25.6] | 22.1  [20.4–23.8] | 25.4  [23.6–27.2] | 25.8  [24.1–27.5] | 27.2  [25.4–28.9] |
| 45-54 | 21.7  [20.2–23.2] | 22.1  [20.5–23.7] | 22.5  [21.0–24.0] | 22.1  [20.5–23.6] | 21.7  [20.2–23.2] | 19.9  [18.4–21.4] | 17.7  [16.3–19.2] | 18.0  [16.7–19.4] | 21.8  [20.3–23.4] | 22.9  [21.4–24.5] | 22.0  [20.4–23.6] |
| 55-64 | 17.9  [16.5–19.3] | 19.0  [17.5–20.4] | 18.3  [16.9–19.7] | 18.1  [16.7–19.5] | 16.9  [15.6–18.3] | 17.4  [16.0–18.9] | 15.7  [14.4–17.1] | 16.0  [14.7–17.4] | 14.8  [13.5–16.2] | 20.6  [19.2–22.1] | 19.6  [18.1–21.1] |
| ≥65 | 10.1  [9.3–11.0] | 10.1  [9.2–11.0] | 10.8  [9.9–11.7] | 9.6  [8.8–10.4] | 10.4  [9.6–11.2] | 9.4  [8.6–10.2] | 9.1  [8.3–9.9] | 10.8  [9.9–11.7] | 10.6  [9.6–11.5] | 11.1  [10.1–12.0] | 11.2  [10.2–12.1] |

### Table S2. Smoking prevalence by age group and year

|  | **Smoking, % [95% confidence interval]** | | | | | | | | | | |
| --- | --- | --- | --- | --- | --- | --- | --- | --- | --- | --- | --- |
|  | **2014** | **2015** | **2016** | **2017** | **2018** | **2019** | **2020** | **2021** | **2022** | **2023** | **2024** |
| 18-24 | 25.3  [23.6–26.9] | 26.5  [24.7–28.3] | 24.9  [23.2–26.7] | 24.1  [22.4–25.9] | 22.3  [20.6–24.1] | 19.2  [17.6–20.8] | 24.2  [22.0–26.3] | 23.6  [21.6–25.7] | 22.1  [20.2–24.0] | 21.9  [19.9–23.8] | 19.9  [18.1–21.8] |
| 25-34 | 25.5  [23.9–27.2] | 24.9  [23.2–26.6] | 23.6  [21.9–25.2] | 23.8  [22.2–25.5] | 24.8  [23.1–26.5] | 23.3  [21.6–24.9] | 24.0  [22.2–25.8] | 25.4  [23.6–27.1] | 24.8  [23.1–26.5] | 22.4  [20.8–24.0] | 21.7  [20.0–23.3] |
| 35-44 | 20.5  [19.0–22.0] | 21.6  [19.9–23.2] | 20.4  [18.8–21.9] | 18.6  [17.1–20.2] | 20.0  [18.4–21.5] | 16.9  [15.4–18.3] | 18.9  [17.2–20.6] | 17.9  [16.4–19.5] | 19.0  [17.4–20.6] | 17.5  [16.1–19.0] | 17.6  [16.1–19.2] |
| 45-54 | 20.2  [18.7–21.7] | 19.6  [18.1–21.1] | 19.8  [18.3–21.2] | 18.9  [17.4–20.3] | 18.4  [17.0–19.8] | 16.7  [15.3–18.1] | 14.2  [12.8–15.5] | 14.3  [13.1–15.6] | 17.3  [15.9–18.8] | 16.5  [15.1–17.9] | 15.7  [14.2–17.1] |
| 55-64 | 16.8  [15.4–18.2] | 17.7  [16.2–19.1] | 16.3  [15.0–17.6] | 15.6  [14.3–16.9] | 15.1  [13.8–16.4] | 14.6  [13.3–15.9] | 13.1  [11.8–14.3] | 12.7  [11.5–13.9] | 12.0  [10.8–13.2] | 16.1  [14.7–17.4] | 14.2  [12.9–15.6] |
| ≥65 | 9.6  [8.7–10.4] | 9.4  [8.5–10.3] | 10.0  [9.1–10.8] | 8.7  [7.9–9.5] | 9.3  [8.5–10.1] | 8.4  [7.6–9.2] | 7.9  [7.2–8.7] | 9.3  [8.5–10.2] | 9.1  [8.2–10.1] | 9.1  [8.3–10.0] | 8.6  [7.8–9.5] |

### Table S3. Vaping prevalence by age group and year

|  | **Vaping, % [95% confidence interval]** | | | | | | | | | | |
| --- | --- | --- | --- | --- | --- | --- | --- | --- | --- | --- | --- |
|  | **2014** | **2015** | **2016** | **2017** | **2018** | **2019** | **2020** | **2021** | **2022** | **2023** | **2024** |
| 18-24 | 5.0  [4.2–5.9] | 6.1  [5.1–7.0] | 6.7  [5.7–7.8] | 5.8  [4.8–6.8] | 5.3  [4.4–6.3] | 5.6  [4.7–6.5] | 8.0  [6.7–9.2] | 11.9  [10.3–13.4] | 18.0  [16.3–19.7] | 24.1  [22.1–26.2] | 25.0  [22.9–27.0] |
| 25-34 | 6.7  [5.7–7.6] | 7.4  [6.4–8.4] | 7.6  [6.6–8.7] | 7.8  [6.8–8.9] | 8.1  [7.0–9.1] | 7.3  [6.3–8.3] | 9.6  [8.4–10.8] | 11.0  [9.8–12.3] | 15.8  [14.4–17.3] | 17.4  [16.0–18.9] | 20.7  [19.1–22.3] |
| 35-44 | 6.0  [5.1–7.0] | 6.9  [5.9–8.0] | 7.4  [6.3–8.4] | 7.0  [6.0–8.0] | 6.8  [5.8–7.8] | 6.4  [5.4–7.4] | 8.1  [7.0–9.3] | 7.7  [6.6–8.8] | 10.6  [9.4–11.9] | 13.9  [12.5–15.3] | 14.2  [12.8–15.6] |
| 45-54 | 6.0  [5.1–6.9] | 7.2  [6.2–8.2] | 6.7  [5.8–7.6] | 6.8  [5.9–7.7] | 6.8  [5.9–7.7] | 6.1  [5.2–7.0] | 6.6  [5.7–7.6] | 6.6  [5.7–7.5] | 8.3  [7.3–9.3] | 10.5  [9.3–11.6] | 10.3  [9.2–11.5] |
| 55-64 | 4.3  [3.5–5.0] | 5.5  [4.6–6.3] | 5.1  [4.3–5.9] | 5.3  [4.5–6.2] | 4.5  [3.8–5.3] | 5.7  [4.8–6.6] | 5.1  [4.3–5.9] | 5.9  [5.0–6.8] | 5.5  [4.6–6.4] | 8.0  [7.1–9.0] | 8.1  [7.1–9.1] |
| ≥65 | 2.2  [1.7–2.6] | 2.1  [1.6–2.5] | 2.1  [1.7–2.5] | 2.1  [1.7–2.5] | 2.2  [1.8–2.6] | 2.0  [1.6–2.4] | 2.2  [1.8–2.6] | 2.5  [2.1–3.0] | 2.3  [1.8–2.7] | 3.1  [2.6–3.6] | 3.9  [3.3–4.5] |

### Table S4. Heated tobacco use prevalence by age group and year

|  | **Heated tobacco use, % [95% confidence interval]** | | | | | | | | | | |
| --- | --- | --- | --- | --- | --- | --- | --- | --- | --- | --- | --- |
|  | **2014** | **2015** | **2016** | **2017** | **2018** | **2019** | **2020** | **2021** | **2022** | **2023** | **2024** |
| 18-24 | - | - | - | 0.2  [0.0–0.3] | 0.0  [0.0–0.0] | 0.1  [0.0–0.3] | 0.1  [0.0–0.2] | 0.6  [0.2–1.0] | 0.9  [0.4–1.4] | 0.4  [0.1–0.7] | 0.4  [0.1–0.8] |
| 25-34 | - | - | - | 0.1  [0.0–0.2] | 0.3  [0.1–0.5] | 0.1  [0.0–0.2] | 0.2  [0.0–0.4] | 0.6  [0.3–0.9] | 0.4  [0.2–0.6] | 0.3  [0.1–0.5] | 0.5  [0.3–0.8] |
| 35-44 | - | - | - | 0.2  [0.1–0.4] | 0.1  [0.0–0.3] | 0.1  [0.0–0.2] | 0.2  [0.0–0.3] | 0.4  [0.1–0.6] | 0.2  [0.0–0.4] | 0.3  [0.1–0.5] | 0.4  [0.2–0.6] |
| 45-54 | - | - | - | 0.1  [0.0–0.2] | 0.1  [0.0–0.2] | 0.2  [0.0–0.4] | 0.3  [0.1–0.5] | 0.2  [0.1–0.3] | 0.3  [0.1–0.5] | 0.3  [0.1–0.5] | 0.1  [0.0–0.2] |
| 55-64 | - | - | - | 0.1  [0.0–0.2] | 0.1  [0.0–0.2] | 0.0  [0.0–0.1] | 0.2  [0.0–0.3] | 0.1  [0.0–0.3] | 0.1  [0.0–0.3] | 0.2  [0.1–0.4] | 0.2  [0.0–0.3] |
| ≥65 | - | - | - | 0.0  [0.0–0.1] | 0.1  [0.0–0.2] | 0.1  [0.0–0.1] | 0.0  [0.0–0.1] | 0.1  [0.0–0.1] | 0.1  [0.0–0.1] | 0.1  [0.0–0.1] | 0.1  [0.0–0.2] |

### Table S5. Nicotine pouch use prevalence by age group and year

|  | **Nicotine pouch use, % [95% confidence interval]** | | | | | | | | | | |
| --- | --- | --- | --- | --- | --- | --- | --- | --- | --- | --- | --- |
|  | **2014** | **2015** | **2016** | **2017** | **2018** | **2019** | **2020** | **2021** | **2022** | **2023** | **2024** |
| 18-24 | - | - | - | - | - | - | - | 0.7  [0.3–1.2] | 0.9  [0.5–1.4] | 1.8  [1.2–2.5] | 3.4  [2.6–4.3] |
| 25-34 | - | - | - | - | - | - | - | 0.5  [0.2–0.7] | 0.6  [0.3–0.8] | 0.8  [0.4–1.2] | 1.3  [0.9–1.7] |
| 35-44 | - | - | - | - | - | - | - | 0.5  [0.2–0.8] | 0.3  [0.1–0.4] | 0.3  [0.1–0.6] | 0.6  [0.3–0.8] |
| 45-54 | - | - | - | - | - | - | - | 0.2  [0.0–0.4] | 0.5  [0.2–0.7] | 0.3  [0.1–0.6] | 0.7  [0.3–1.0] |
| 55-64 | - | - | - | - | - | - | - | 0.1  [0.0–0.3] | 0.1  [0.0–0.2] | 0.2  [0.1–0.4] | 0.1  [0.0–0.3] |
| ≥65 | - | - | - | - | - | - | - | 0.0  [0.0–0.1] | 0.0  [0.0–0.1] | 0.1  [0.0–0.2] | 0.1  [0.0–0.2] |
