## Supplementary file 2 for "The changing face of nicotine use in England: age-specific annual trends, 2014-2024"

### Table S6. Exclusive smoking prevalence among adults who smoke or vape, by age group and year

|  | **Exclusive smoking, % [95% confidence interval]** | | | | | | | | | | |
| --- | --- | --- | --- | --- | --- | --- | --- | --- | --- | --- | --- |
|  | **2014** | **2015** | **2016** | **2017** | **2018** | **2019** | **2020** | **2021** | **2022** | **2023** | **2024** |
| 18-24 | 80.7  [77.8–83.6] | 77.9  [74.7–81.1] | 74.3  [70.9–77.8] | 77.4  [74.0–80.9] | 77.6  [74.0–81.2] | 73.0  [69.1–77.0] | 70.8  [66.5–75.0] | 58.4  [53.9–62.9] | 40.6  [36.6–44.6] | 29.3  [25.5–33.1] | 29.6  [25.9–33.2] |
| 25-34 | 75.3  [72.2–78.4] | 72.7  [69.4–76.0] | 70.9  [67.4–74.4] | 71.3  [67.8–74.7] | 71.0  [67.7–74.3] | 72.3  [68.9–75.7] | 66.4  [62.7–70.1] | 63.4  [59.9–67.0] | 51.5  [48.1–55.0] | 45.1  [41.6–48.5] | 39.5  [36.2–42.9] |
| 35-44 | 73.1  [69.5–76.7] | 70.9  [67.2–74.6] | 68.6  [64.7–72.4] | 67.7  [63.9–71.6] | 70.3  [66.6–74.0] | 67.8  [63.6–72.0] | 65.7  [61.5–69.9] | 64.7  [60.5–68.8] | 58.0  [53.9–62.1] | 45.6  [41.7–49.4] | 46.9  [43.0–50.8] |
| 45-54 | 72.3  [68.7–75.8] | 67.3  [63.5–71.1] | 70.2  [66.7–73.6] | 69.2  [65.5–72.8] | 68.6  [64.9–72.2] | 69.0  [65.0–73.0] | 62.2  [57.8–66.7] | 63.3  [59.3–67.4] | 61.7  [57.7–65.7] | 54.2  [50.4–58.1] | 52.4  [48.2–56.7] |
| 55-64 | 76.2  [72.3–80.1] | 71.2  [67.2–75.1] | 72.1  [68.3–75.9] | 70.4  [66.5–74.3] | 73.2  [69.4–77.0] | 67.3  [63.1–71.5] | 67.9  [63.5–72.3] | 63.1  [58.7–67.5] | 62.6  [57.8–67.5] | 60.5  [56.6–64.5] | 58.5  [54.2–62.8] |
| ≥65 | 78.5  [74.7–82.3] | 79.4  [75.4–83.4] | 80.5  [77.2–83.8] | 78.4  [74.8–82.0] | 78.5  [75.0–81.9] | 78.6  [74.9–82.4] | 75.7  [71.6–79.8] | 76.7  [72.9–80.4] | 78.5  [74.6–82.5] | 71.8  [67.7–75.9] | 65.2  [60.6–69.7] |

### Table S7. Exclusive vaping prevalence among adults who smoke or vape, by age group and year

|  | **Exclusive vaping, % [95% confidence interval]** | | | | | | | | | | |
| --- | --- | --- | --- | --- | --- | --- | --- | --- | --- | --- | --- |
|  | **2014** | **2015** | **2016** | **2017** | **2018** | **2019** | **2020** | **2021** | **2022** | **2023** | **2024** |
| 18-24 | 3.0  [1.7–4.3] | 3.9  [2.2–5.5] | 5.0  [3.3–6.7] | 5.7  [3.8–7.6] | 6.1  [4.0–8.2] | 7.7  [5.1–10.2] | 11.1  [8.3–13.9] | 17.0  [13.6–20.4] | 27.0  [23.4–30.6] | 36.0  [32.0–39.9] | 43.7  [39.8–47.7] |
| 25-34 | 5.6  [3.9–7.3] | 7.9  [5.8–10.0] | 10.0  [7.7–12.4] | 12.3  [9.8–14.9] | 11.0  [8.7–13.3] | 11.5  [9.0–14.1] | 16.0  [13.2–18.7] | 15.9  [13.2–18.5] | 24.0  [21.1–27.0] | 29.4  [26.3–32.6] | 36.6  [33.3–39.9] |
| 35-44 | 8.9  [6.5–11.2] | 9.6  [7.1–12.0] | 13.0  [10.1–15.8] | 13.8  [10.9–16.8] | 12.8  [10.0–15.6] | 15.0  [11.7–18.3] | 20.3  [16.8–23.8] | 18.0  [14.7–21.4] | 25.1  [21.5–28.6] | 31.3  [27.7–34.9] | 34.1  [30.5–37.7] |
| 45-54 | 6.8  [4.8–8.9] | 11.2  [8.5–13.8] | 12.2  [9.7–14.8] | 14.3  [11.6–17.1] | 15.0  [12.1–18.0] | 15.5  [12.5–18.5] | 19.2  [15.7–22.7] | 19.9  [16.5–23.2] | 20.2  [16.9–23.5] | 27.7  [24.2–31.2] | 27.9  [24.1–31.6] |
| 55-64 | 6.3  [4.0–8.6] | 6.8  [4.6–9.0] | 11.0  [8.3–13.8] | 13.4  [10.5–16.3] | 10.7  [8.1–13.3] | 16.4  [13.0–19.8] | 17.1  [13.6–20.6] | 20.5  [16.8–24.2] | 18.7  [14.8–22.5] | 21.2  [18.0–24.5] | 27.2  [23.4–31.0] |
| ≥65 | 5.7  [3.6–7.7] | 6.9  [4.4–9.4] | 7.8  [5.6–10.0] | 9.7  [7.1–12.3] | 10.4  [7.8–12.9] | 10.4  [7.7–13.2] | 12.7  [9.5–15.9] | 12.8  [9.8–15.8] | 13.5  [10.2–16.8] | 17.2  [13.7–20.6] | 22.5  [18.5–26.4] |

### Table S8. Dual use prevalence among adults who smoke or vape, by age group and year

|  | **Dual use of smoking and vaping, % [95% confidence interval]** | | | | | | | | | | |
| --- | --- | --- | --- | --- | --- | --- | --- | --- | --- | --- | --- |
|  | **2014** | **2015** | **2016** | **2017** | **2018** | **2019** | **2020** | **2021** | **2022** | **2023** | **2024** |
| 18-24 | 16.3  [13.6–19.0] | 18.2  [15.3–21.1] | 20.7  [17.5–23.9] | 16.9  [13.8–20.0] | 16.3  [13.2–19.5] | 19.3  [15.9–22.7] | 18.2  [14.5–21.8] | 24.6  [20.7–28.6] | 32.4  [28.6–36.2] | 34.8  [30.9–38.6] | 26.7  [23.2–30.1] |
| 25-34 | 19.1  [16.3–21.9] | 19.4  [16.5–22.3] | 19.1  [16.0–22.1] | 16.4  [13.6–19.2] | 18.0  [15.2–20.7] | 16.2  [13.5–18.9] | 17.7  [14.6–20.7] | 20.7  [17.7–23.7] | 24.4  [21.4–27.4] | 25.5  [22.4–28.6] | 23.9  [20.8–26.9] |
| 35-44 | 18.0  [14.9–21.1] | 19.6  [16.3–22.8] | 18.5  [15.3–21.6] | 18.4  [15.3–21.6] | 16.9  [13.9–19.9] | 17.2  [13.8–20.6] | 14.1  [10.9–17.2] | 17.3  [14.0–20.5] | 17.0  [14.0–19.9] | 23.1  [19.8–26.4] | 18.9  [15.9–22.0] |
| 45-54 | 20.9  [17.7–24.1] | 21.6  [18.2–24.9] | 17.6  [14.8–20.5] | 16.5  [13.6–19.4] | 16.4  [13.6–19.2] | 15.5  [12.3–18.8] | 18.5  [14.9–22.2] | 16.8  [13.6–20.0] | 18.1  [14.9–21.3] | 18.0  [15.1–21.0] | 19.7  [16.3–23.1] |
| 55-64 | 17.5  [14.1–21.0] | 22.1  [18.4–25.7] | 16.9  [13.7–20.0] | 16.2  [13.0–19.4] | 16.0  [12.9–19.2] | 16.3  [13.0–19.6] | 15.0  [11.7–18.3] | 16.4  [12.9–19.8] | 18.7  [14.6–22.8] | 18.2  [15.1–21.3] | 14.3  [11.2–17.4] |
| ≥65 | 15.8  [12.4–19.2] | 13.7  [10.3–17.2] | 11.7  [9.0–14.4] | 11.9  [9.1–14.8] | 11.1  [8.5–13.8] | 10.9  [8.0–13.8] | 11.6  [8.6–14.7] | 10.5  [7.9–13.2] | 8.0  [5.5–10.6] | 11.0  [8.1–13.9] | 12.4  [9.2–15.5] |

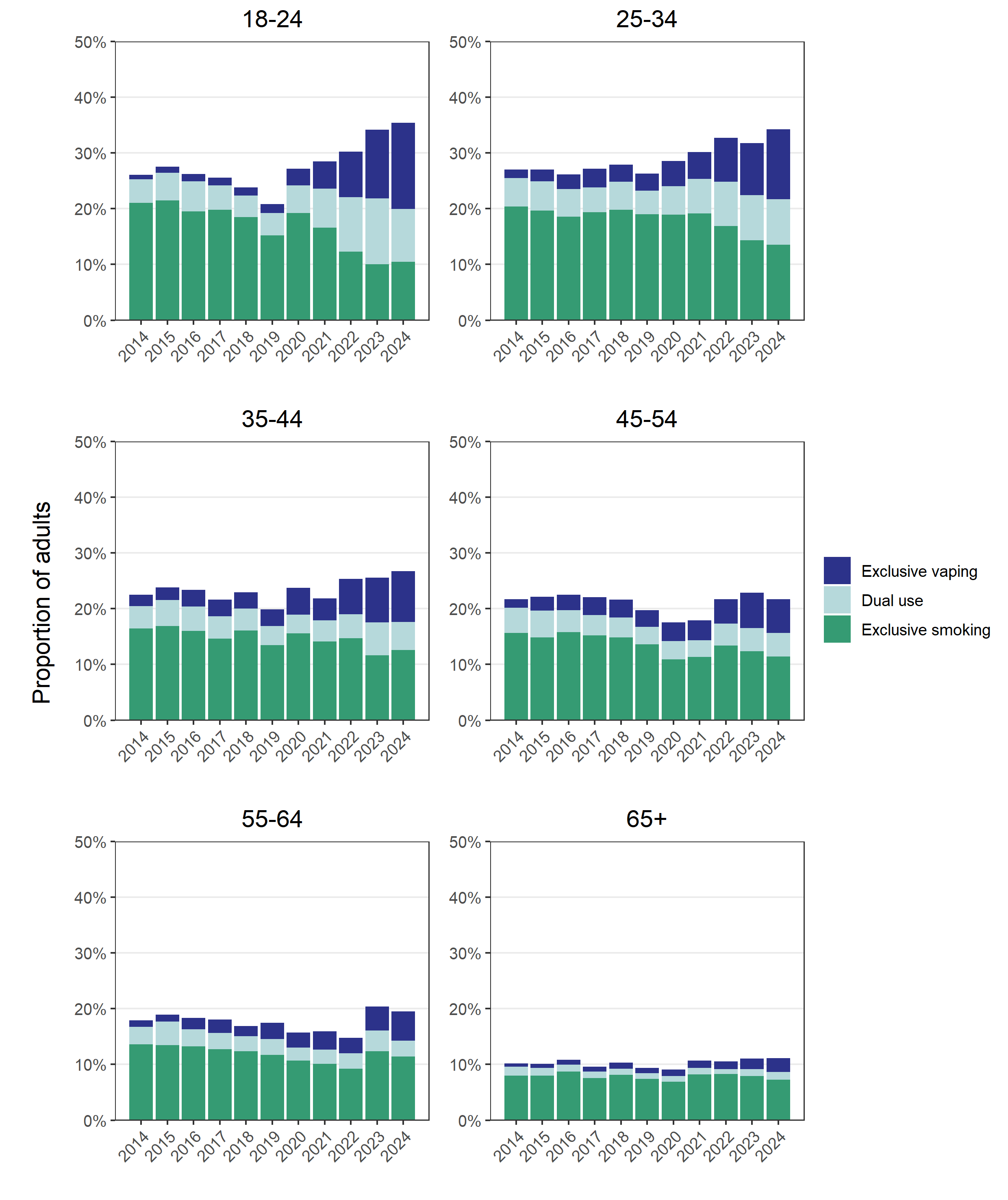

### Figure S1. Prevalence of exclusive smoking, exclusive vaping, and dual use among adults in England, 2014 to 2024. Estimates with 95% confidence intervals are provided in Tables S9-11.

### Table S9. Exclusive smoking prevalence among adults, by age group and year

|  | **Exclusive smoking, % [95% confidence interval]** | | | | | | | | | | |
| --- | --- | --- | --- | --- | --- | --- | --- | --- | --- | --- | --- |
|  | **2014** | **2015** | **2016** | **2017** | **2018** | **2019** | **2020** | **2021** | **2022** | **2023** | **2024** |
| 18-24 | 21.0  [19.5–22.6] | 21.5  [19.8–23.1] | 19.5  [17.9–21.1] | 19.8  [18.2–21.5] | 18.5  [16.9–20.0] | 15.2  [13.8–16.7] | 19.2  [17.3–21.2] | 16.6  [14.8–18.4] | 12.3  [10.8–13.8] | 10.0  [8.5–11.5] | 10.5  [9.0–11.9] |
| 25-34 | 20.4  [18.9–21.9] | 19.7  [18.1–21.2] | 18.6  [17.1–20.0] | 19.4  [17.8–21.0] | 19.8  [18.2–21.4] | 19.0  [17.5–20.5] | 19.0  [17.3–20.6] | 19.1  [17.5–20.7] | 16.9  [15.4–18.3] | 14.3  [12.9–15.7] | 13.5  [12.2–14.9] |
| 35-44 | 16.4  [15.0–17.8] | 16.9  [15.4–18.4] | 16.0  [14.6–17.5] | 14.7  [13.3–16.0] | 16.1  [14.7–17.5] | 13.5  [12.1–14.8] | 15.6  [14.0–17.1] | 14.1  [12.7–15.5] | 14.7  [13.2–16.2] | 11.6  [10.4–12.9] | 12.6  [11.2–13.9] |
| 45-54 | 15.7  [14.3–17.0] | 14.9  [13.6–16.2] | 15.8  [14.5–17.1] | 15.2  [13.9–16.6] | 14.8  [13.6–16.1] | 13.6  [12.3–15.0] | 10.9  [9.7–12.1] | 11.3  [10.2–12.4] | 13.4  [12.1–14.7] | 12.4  [11.2–13.6] | 11.4  [10.1–12.7] |
| 55-64 | 13.6  [12.4–14.9] | 13.5  [12.2–14.8] | 13.2  [12.0–14.4] | 12.7  [11.5–13.9] | 12.4  [11.2–13.6] | 11.7  [10.5–12.9] | 10.7  [9.5–11.9] | 10.1  [9.0–11.2] | 9.2  [8.1–10.3] | 12.4  [11.2–13.5] | 11.4  [10.2–12.7] |
| ≥65 | 8.0  [7.2–8.8] | 8.0  [7.2–8.8] | 8.7  [7.9–9.5] | 7.5  [6.8–8.3] | 8.1  [7.4–8.9] | 7.4  [6.7–8.1] | 6.9  [6.2–7.6] | 8.2  [7.4–9.0] | 8.3  [7.4–9.2] | 7.9  [7.1–8.7] | 7.3  [6.5–8.1] |

### Table S10. Exclusive vaping prevalence among adults, by age group and year

|  | **Exclusive vaping, % [95% confidence interval]** | | | | | | | | | | |
| --- | --- | --- | --- | --- | --- | --- | --- | --- | --- | --- | --- |
|  | **2014** | **2015** | **2016** | **2017** | **2018** | **2019** | **2020** | **2021** | **2022** | **2023** | **2024** |
| 18-24 | 0.8  [0.4–1.1] | 1.1  [0.6–1.5] | 1.3  [0.9–1.8] | 1.4  [1.0–1.9] | 1.4  [0.9–2.0] | 1.6  [1.0–2.2] | 3.0  [2.2–3.8] | 4.8  [3.8–5.9] | 8.2  [6.9–9.4] | 12.3  [10.7–13.9] | 15.5  [13.8–17.2] |
| 25-34 | 1.5  [1.0–2.0] | 2.1  [1.5–2.7] | 2.6  [2.0–3.3] | 3.4  [2.6–4.1] | 3.1  [2.4–3.7] | 3.0  [2.3–3.7] | 4.6  [3.7–5.4] | 4.8  [3.9–5.6] | 7.9  [6.8–8.9] | 9.3  [8.2–10.5] | 12.5  [11.2–13.8] |
| 35-44 | 2.0  [1.4–2.5] | 2.3  [1.7–2.9] | 3.0  [2.3–3.7] | 3.0  [2.3–3.7] | 2.9  [2.3–3.6] | 3.0  [2.3–3.7] | 4.8  [3.9–5.7] | 3.9  [3.1–4.7] | 6.3  [5.3–7.4] | 8.0  [6.9–9.1] | 9.1  [8.0–10.2] |
| 45-54 | 1.5  [1.0–1.9] | 2.5  [1.9–3.1] | 2.8  [2.2–3.4] | 3.2  [2.5–3.8] | 3.3  [2.6–3.9] | 3.1  [2.4–3.7] | 3.4  [2.7–4.0] | 3.6  [2.9–4.2] | 4.4  [3.6–5.2] | 6.3  [5.4–7.2] | 6.1  [5.1–7.0] |
| 55-64 | 1.1  [0.7–1.5] | 1.3  [0.9–1.7] | 2.0  [1.5–2.5] | 2.4  [1.9–3.0] | 1.8  [1.4–2.3] | 2.9  [2.2–3.5] | 2.7  [2.1–3.3] | 3.3  [2.6–3.9] | 2.8  [2.1–3.4] | 4.3  [3.6–5.1] | 5.3  [4.5–6.2] |
| ≥65 | 0.6  [0.4–0.8] | 0.7  [0.4–1.0] | 0.8  [0.6–1.1] | 0.9  [0.7–1.2] | 1.1  [0.8–1.3] | 1.0  [0.7–1.3] | 1.2  [0.8–1.5] | 1.4  [1.0–1.7] | 1.4  [1.1–1.8] | 1.9  [1.5–2.3] | 2.5  [2.0–3.0] |

### Table S11. Dual use prevalence among adults, by age group and year

|  | **Dual use of smoking and vaping, % [95% confidence interval]** | | | | | | | | | | |
| --- | --- | --- | --- | --- | --- | --- | --- | --- | --- | --- | --- |
|  | **2014** | **2015** | **2016** | **2017** | **2018** | **2019** | **2020** | **2021** | **2022** | **2023** | **2024** |
| 18-24 | 4.3  [3.5–5.0] | 5.0  [4.1–5.9] | 5.4  [4.5–6.3] | 4.3  [3.5–5.2] | 3.9  [3.1–4.7] | 4.0  [3.3–4.8] | 4.9  [3.9–6.0] | 7.0  [5.8–8.3] | 9.8  [8.5–11.1] | 11.9  [10.3–13.4] | 9.5  [8.1–10.8] |
| 25-34 | 5.2  [4.3–6.0] | 5.2  [4.4–6.1] | 5.0  [4.1–5.9] | 4.5  [3.7–5.3] | 5.0  [4.2–5.8] | 4.3  [3.5–5.0] | 5.1  [4.1–6.0] | 6.2  [5.2–7.2] | 8.0  [6.9–9.1] | 8.1  [7.0–9.2] | 8.2  [7.0–9.3] |
| 35-44 | 4.1  [3.3–4.8] | 4.7  [3.8–5.5] | 4.3  [3.5–5.1] | 4.0  [3.3–4.7] | 3.9  [3.1–4.6] | 3.4  [2.7–4.1] | 3.3  [2.5–4.1] | 3.8  [3.0–4.5] | 4.3  [3.5–5.1] | 5.9  [5.0–6.8] | 5.1  [4.2–6.0] |
| 45-54 | 4.5  [3.8–5.3] | 4.8  [3.9–5.6] | 4.0  [3.3–4.7] | 3.6  [3.0–4.3] | 3.6  [2.9–4.2] | 3.1  [2.4–3.8] | 3.3  [2.5–4.0] | 3.0  [2.4–3.6] | 3.9  [3.2–4.7] | 4.1  [3.4–4.8] | 4.3  [3.5–5.1] |
| 55-64 | 3.1  [2.5–3.8] | 4.2  [3.4–5.0] | 3.1  [2.5–3.7] | 2.9  [2.3–3.5] | 2.7  [2.1–3.3] | 2.8  [2.2–3.5] | 2.4  [1.8–2.9] | 2.6  [2.0–3.2] | 2.8  [2.1–3.4] | 3.7  [3.0–4.4] | 2.8  [2.2–3.4] |
| ≥65 | 1.6  [1.2–2.0] | 1.4  [1.0–1.8] | 1.3  [1.0–1.6] | 1.1  [0.9–1.4] | 1.2  [0.9–1.4] | 1.0  [0.7–1.3] | 1.1  [0.8–1.4] | 1.1  [0.8–1.4] | 0.8  [0.6–1.1] | 1.2  [0.9–1.6] | 1.4  [1.0–1.8] |
