## Supplementary file 3 for "The changing face of nicotine use in England: age-specific annual trends, 2014-2024"

### Table S12. Proportion who smoke cigarettes daily among adults who smoke, by age group and year

|  | **Daily cigarette smoking, % [95% confidence interval]** | | | | | | | | | | |
| --- | --- | --- | --- | --- | --- | --- | --- | --- | --- | --- | --- |
|  | **2014** | **2015** | **2016** | **2017** | **2018** | **2019** | **2020** | **2021** | **2022** | **2023** | **2024** |
| 18-24 | 84.5  [81.7–87.2] | 84.8  [82.0–87.5] | 80.7  [77.6–83.8] | 76.6  [73.1–80.1] | 72.8  [69.0–76.6] | 74.0  [70.1–77.9] | 64.1  [59.1–69.0] | 54.7  [49.7–59.7] | 53.6  [48.8–58.3] | 44.8  [39.7–49.9] | 46.7  [41.5–52.0] |
| 25-34 | 86.9  [84.4–89.5] | 85.0  [82.1–87.9] | 84.8  [82.1–87.6] | 82.6  [79.6–85.7] | 80.1  [77.0–83.2] | 84.0  [81.1–87.0] | 72.4  [68.5–76.3] | 64.4  [60.5–68.2] | 61.9  [58.1–65.8] | 61.9  [57.9–65.9] | 63.4  [59.3–67.6] |
| 35-44 | 85.6  [82.6–88.5] | 84.9  [81.7–88.2] | 88.7  [85.9–91.4] | 86.7  [83.6–89.8] | 84.1  [81.0–87.3] | 84.8  [81.4–88.2] | 70.1  [65.5–74.7] | 71.7  [67.4–75.9] | 71.7  [67.5–75.9] | 68.7  [64.5–72.8] | 64.4  [59.8–69.0] |
| 45-54 | 88.1  [85.4–90.8] | 86.4  [83.3–89.4] | 88.1  [85.5–90.7] | 87.8  [85.1–90.6] | 84.7  [81.7–87.7] | 87.6  [84.6–90.7] | 72.9  [68.4–77.5] | 78.0  [74.2–81.8] | 74.2  [70.3–78.2] | 75.6  [71.8–79.3] | 74.2  [69.8–78.7] |
| 55-64 | 87.5  [84.4–90.6] | 88.0  [84.9–91.0] | 87.9  [85.0–90.9] | 86.7  [83.6–89.8] | 89.7  [86.9–92.6] | 90.2  [87.3–93.1] | 80.0  [75.7–84.2] | 79.7  [75.6–83.9] | 70.7  [65.7–75.7] | 76.7  [73.0–80.5] | 77.7  [73.5–81.8] |
| ≥65 | 87.9  [84.7–91.0] | 87.0  [83.8–90.3] | 85.9  [82.9–89.0] | 87.0  [83.9–90.0] | 83.4  [80.1–86.7] | 87.6  [84.4–90.8] | 80.2  [76.2–84.2] | 76.4  [72.4–80.4] | 79.3  [75.1–83.6] | 80.7  [76.9–84.6] | 76.1  [71.8–80.4] |

### Table S13. Proportion who smoke cigarettes non-daily among adults who smoke, by age group and year

|  | **Non-daily cigarette smoking, % [95% confidence interval]** | | | | | | | | | | |
| --- | --- | --- | --- | --- | --- | --- | --- | --- | --- | --- | --- |
|  | **2014** | **2015** | **2016** | **2017** | **2018** | **2019** | **2020** | **2021** | **2022** | **2023** | **2024** |
| 18-24 | 14.5  [11.8–17.2] | 12.7  [10.2–15.2] | 17.1  [14.2–20.1] | 21.3  [17.9–24.6] | 25.7  [22.0–29.5] | 22.5  [18.9–26.1] | 28.8  [24.1–33.5] | 34.4  [29.6–39.2] | 33.9  [29.4–38.4] | 40.8  [35.8–45.8] | 43.3  [38.1–48.5] |
| 25-34 | 11.9  [9.4–14.3] | 13.9  [11.1–16.8] | 13.0  [10.4–15.6] | 15.6  [12.6–18.5] | 18.3  [15.3–21.4] | 13.9  [11.2–16.7] | 18.7  [15.4–22.1] | 25.8  [22.3–29.4] | 26.9  [23.3–30.4] | 24.7  [21.2–28.3] | 26.5  [22.8–30.3] |
| 35-44 | 12.6  [9.8–15.5] | 12.8  [9.8–15.8] | 9.2  [6.8–11.6] | 12.2  [9.2–15.2] | 14.4  [11.3–17.4] | 12.6  [9.6–15.7] | 21.8  [17.7–25.9] | 20.1  [16.3–24.0] | 17.1  [13.7–20.5] | 21.1  [17.5–24.7] | 26.6  [22.4–30.9] |
| 45-54 | 9.2  [6.7–11.6] | 11.2  [8.3–14.0] | 9.8  [7.4–12.1] | 10.7  [8.2–13.3] | 13.3  [10.4–16.2] | 10.7  [7.8–13.5] | 18.3  [14.3–22.3] | 12.8  [9.8–15.8] | 15.2  [11.9–18.4] | 15.6  [12.5–18.7] | 17.4  [13.6–21.3] |
| 55-64 | 10.1  [7.2–13.0] | 9.3  [6.6–12.0] | 9.3  [6.7–11.9] | 10.9  [8.1–13.7] | 7.5  [5.1–10.0] | 6.3  [3.9–8.7] | 14.5  [10.7–18.2] | 12.8  [9.3–16.3] | 17.1  [13.1–21.1] | 13.7  [10.6–16.7] | 15.0  [11.4–18.6] |
| ≥65 | 6.5  [4.0–9.0] | 8.5  [5.7–11.3] | 7.9  [5.5–10.4] | 7.9  [5.4–10.4] | 10.0  [7.3–12.7] | 7.4  [4.9–10.0] | 8.8  [6.0–11.5] | 9.8  [6.8–12.7] | 9.0  [5.9–12.1] | 9.2  [6.4–12.0] | 14.6  [10.9–18.3] |

### Table S14. Proportion who exclusively smoke non-cigarette tobacco among adults who smoke, by age group and year

|  | **Exclusive non-cigarette smoking, % [95% confidence interval]** | | | | | | | | | | |
| --- | --- | --- | --- | --- | --- | --- | --- | --- | --- | --- | --- |
|  | **2014** | **2015** | **2016** | **2017** | **2018** | **2019** | **2020** | **2021** | **2022** | **2023** | **2024** |
| 18-24 | 1.0  [0.3–1.8] | 2.5  [1.3–3.8] | 2.2  [1.0–3.4] | 2.1  [0.9–3.3] | 1.5  [0.5–2.4] | 3.5  [1.6–5.3] | 7.2  [4.6–9.7] | 10.9  [7.9–14.0] | 12.5  [9.5–15.6] | 14.4  [10.8–17.9] | 9.9  [7.0–12.9] |
| 25-34 | 1.2  [0.4–2.0] | 1.1  [0.4–1.8] | 2.1  [1.1–3.2] | 1.8  [0.7–2.9] | 1.6  [0.6–2.5] | 2.0  [0.9–3.1] | 8.9  [6.3–11.4] | 9.8  [7.5–12.2] | 11.2  [8.8–13.7] | 13.4  [10.7–16.1] | 10.0  [7.4–12.6] |
| 35-44 | 1.8  [0.7–2.9] | 2.2  [0.8–3.7] | 2.1  [0.8–3.5] | 1.1  [0.2–2.0] | 1.5  [0.5–2.5] | 2.6  [0.9–4.2] | 8.1  [5.2–11.0] | 8.2  [5.8–10.7] | 11.2  [8.2–14.1] | 10.2  [7.5–12.9] | 9.0  [6.4–11.5] |
| 45-54 | 2.7  [1.4–4.1] | 2.4  [1.1–3.8] | 2.1  [0.9–3.3] | 1.4  [0.4–2.4] | 2.0  [0.9–3.1] | 1.7  [0.5–3.0] | 8.8  [5.9–11.7] | 9.2  [6.5–11.9] | 10.6  [7.9–13.3] | 8.9  [6.3–11.4] | 8.3  [5.5–11.1] |
| 55-64 | 2.4  [1.0–3.7] | 2.7  [1.1–4.4] | 2.8  [1.3–4.2] | 2.4  [0.9–3.9] | 2.7  [1.3–4.2] | 3.5  [1.6–5.3] | 5.5  [3.1–8.0] | 7.4  [4.8–10.0] | 12.2  [8.4–15.9] | 9.6  [7.1–12.1] | 7.4  [4.9–9.8] |
| ≥65 | 5.6  [3.6–7.6] | 4.4  [2.5–6.4] | 6.1  [4.1–8.2] | 5.1  [3.2–7.0] | 6.6  [4.4–8.7] | 5.0  [2.9–7.0] | 11.0  [7.8–14.2] | 13.8  [10.7–17.0] | 11.7  [8.4–15.0] | 10.1  [7.1–13.0] | 9.3  [6.6–12.0] |

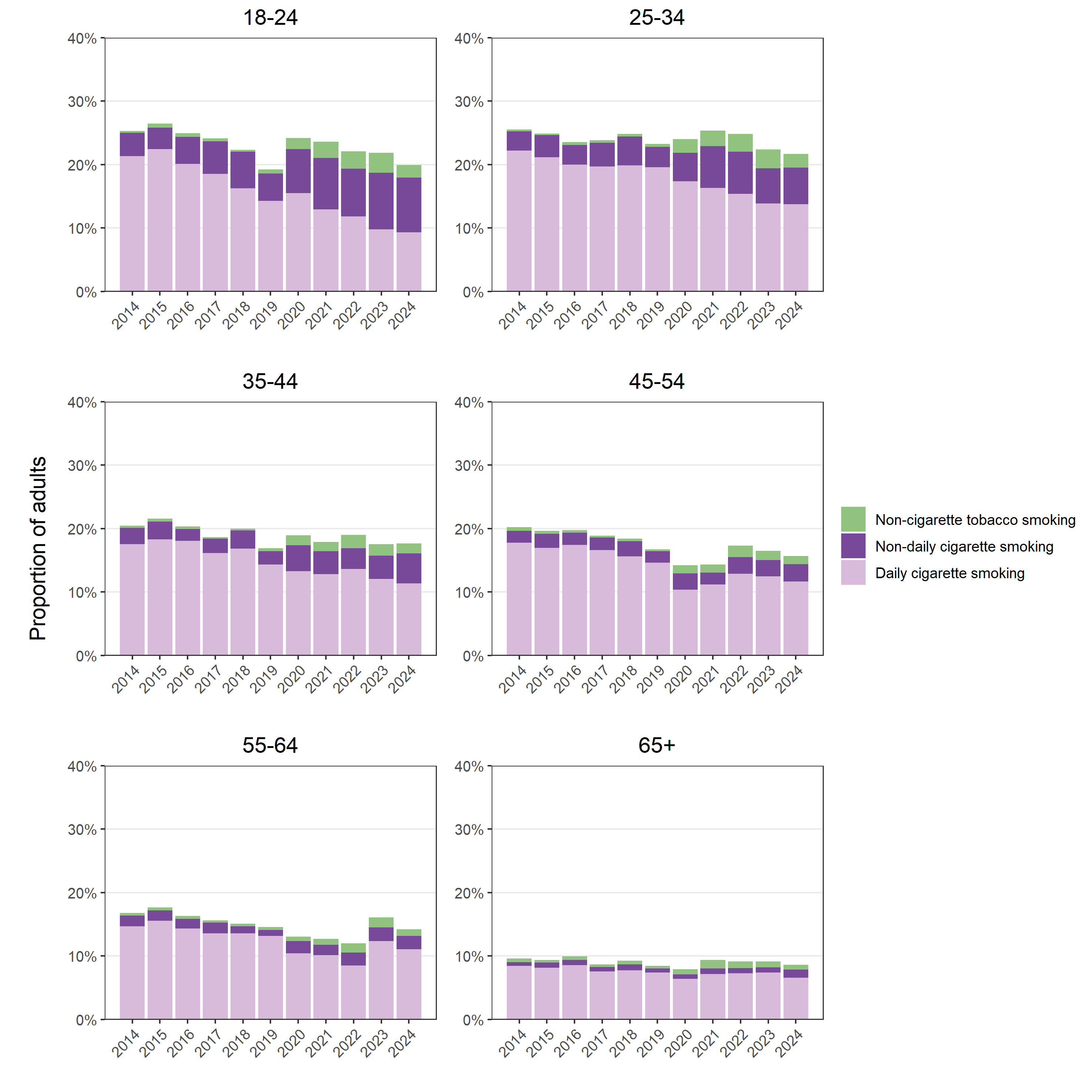

### Figure S2. Smoking type and pattern among adults in England, 2014 to 2024. Estimates with 95% confidence intervals are provided in Tables S15-17.

### Table S15. Proportion who smoke cigarettes daily among adults, by age group and year

|  | **Daily cigarette smoking, % [95% confidence interval]** | | | | | | | | | | |
| --- | --- | --- | --- | --- | --- | --- | --- | --- | --- | --- | --- |
|  | **2014** | **2015** | **2016** | **2017** | **2018** | **2019** | **2020** | **2021** | **2022** | **2023** | **2024** |
| 18-24 | 21.4  [19.8–22.9] | 22.4  [20.8–24.1] | 20.1  [18.5–21.7] | 18.5  [16.9–20.1] | 16.3  [14.7–17.8] | 14.2  [12.8–15.7] | 15.5  [13.7–17.3] | 12.9  [11.3–14.5] | 11.8  [10.4–13.3] | 9.8  [8.4–11.2] | 9.3  [7.9–10.7] |
| 25-34 | 22.2  [20.6–23.7] | 21.2  [19.6–22.7] | 20.0  [18.5–21.5] | 19.7  [18.1–21.3] | 19.9  [18.3–21.4] | 19.6  [18.0–21.1] | 17.4  [15.8–19.0] | 16.3  [14.8–17.8] | 15.4  [14.0–16.8] | 13.9  [12.5–15.2] | 13.8  [12.3–15.2] |
| 35-44 | 17.5  [16.1–19.0] | 18.3  [16.8–19.8] | 18.1  [16.6–19.5] | 16.2  [14.7–17.6] | 16.8  [15.4–18.3] | 14.3  [12.9–15.7] | 13.3  [11.8–14.7] | 12.8  [11.5–14.2] | 13.6  [12.2–15.0] | 12.0  [10.8–13.3] | 11.4  [10.0–12.7] |
| 45-54 | 17.8  [16.4–19.2] | 17.0  [15.6–18.4] | 17.4  [16.0–18.8] | 16.6  [15.2–17.9] | 15.6  [14.3–16.9] | 14.6  [13.3–16.0] | 10.3  [9.2–11.5] | 11.2  [10.1–12.3] | 12.9  [11.5–14.2] | 12.5  [11.2–13.7] | 11.6  [10.4–12.9] |
| 55-64 | 14.7  [13.4–16.0] | 15.5  [14.2–16.9] | 14.3  [13.1–15.6] | 13.6  [12.3–14.8] | 13.5  [12.3–14.8] | 13.2  [11.9–14.4] | 10.4  [9.3–11.6] | 10.1  [9.0–11.2] | 8.5  [7.4–9.5] | 12.3  [11.1–13.5] | 11.1  [9.8–12.3] |
| ≥65 | 8.4  [7.6–9.2] | 8.2  [7.4–9.0] | 8.6  [7.8–9.3] | 7.6  [6.8–8.3] | 7.7  [7.0–8.5] | 7.4  [6.6–8.1] | 6.4  [5.7–7.1] | 7.1  [6.4–7.9] | 7.2  [6.4–8.1] | 7.4  [6.6–8.2] | 6.6  [5.8–7.4] |

### Table S16. Proportion who smoke cigarettes non-daily among adults, by age group and year

|  | **Non-daily cigarette smoking, % [95% confidence interval]** | | | | | | | | | | |
| --- | --- | --- | --- | --- | --- | --- | --- | --- | --- | --- | --- |
|  | **2014** | **2015** | **2016** | **2017** | **2018** | **2019** | **2020** | **2021** | **2022** | **2023** | **2024** |
| 18-24 | 3.7  [3.0–4.4] | 3.4  [2.7–4.1] | 4.3  [3.5–5.1] | 5.1  [4.3–6.0] | 5.7  [4.8–6.7] | 4.3  [3.6–5.1] | 7.0  [5.7–8.3] | 8.1  [6.8–9.5] | 7.5  [6.3–8.7] | 8.9  [7.6–10.3] | 8.6  [7.3–10.0] |
| 25-34 | 3.0  [2.4–3.7] | 3.5  [2.7–4.2] | 3.1  [2.4–3.7] | 3.7  [3.0–4.5] | 4.5  [3.7–5.4] | 3.2  [2.6–3.9] | 4.5  [3.6–5.4] | 6.6  [5.5–7.6] | 6.7  [5.7–7.7] | 5.5  [4.7–6.4] | 5.8  [4.9–6.7] |
| 35-44 | 2.6  [2.0–3.2] | 2.8  [2.1–3.4] | 1.9  [1.4–2.4] | 2.3  [1.7–2.9] | 2.9  [2.2–3.5] | 2.1  [1.6–2.7] | 4.1  [3.3–5.0] | 3.6  [2.8–4.4] | 3.3  [2.6–3.9] | 3.7  [3.0–4.4] | 4.7  [3.9–5.5] |
| 45-54 | 1.8  [1.3–2.4] | 2.2  [1.6–2.8] | 1.9  [1.4–2.4] | 2.0  [1.5–2.5] | 2.4  [1.9–3.0] | 1.8  [1.3–2.3] | 2.6  [2.0–3.2] | 1.8  [1.4–2.3] | 2.6  [2.0–3.2] | 2.6  [2.0–3.1] | 2.7  [2.1–3.4] |
| 55-64 | 1.7  [1.2–2.2] | 1.6  [1.2–2.1] | 1.5  [1.1–2.0] | 1.7  [1.2–2.2] | 1.1  [0.7–1.5] | 0.9  [0.6–1.3] | 1.9  [1.4–2.4] | 1.6  [1.2–2.1] | 2.1  [1.5–2.6] | 2.2  [1.7–2.7] | 2.1  [1.6–2.7] |
| ≥65 | 0.6  [0.4–0.9] | 0.8  [0.5–1.1] | 0.8  [0.5–1.0] | 0.7  [0.5–0.9] | 0.9  [0.7–1.2] | 0.6  [0.4–0.8] | 0.7  [0.5–0.9] | 0.9  [0.6–1.2] | 0.8  [0.5–1.1] | 0.8  [0.6–1.1] | 1.3  [0.9–1.6] |

### Table S17. Proportion who exclusively smoke non-cigarette tobacco among adults, by age group and year

|  | **Exclusive non-cigarette smoking, % [95% confidence interval]** | | | | | | | | | | |
| --- | --- | --- | --- | --- | --- | --- | --- | --- | --- | --- | --- |
|  | **2014** | **2015** | **2016** | **2017** | **2018** | **2019** | **2020** | **2021** | **2022** | **2023** | **2024** |
| 18-24 | 0.3  [0.1–0.4] | 0.7  [0.3–1.0] | 0.5  [0.3–0.8] | 0.5  [0.2–0.8] | 0.3  [0.1–0.5] | 0.7  [0.3–1.0] | 1.7  [1.1–2.4] | 2.6  [1.8–3.3] | 2.8  [2.1–3.5] | 3.1  [2.3–4.0] | 2.0  [1.4–2.6] |
| 25-34 | 0.3  [0.1–0.5] | 0.3  [0.1–0.4] | 0.5  [0.2–0.8] | 0.4  [0.2–0.7] | 0.4  [0.2–0.6] | 0.5  [0.2–0.7] | 2.1  [1.5–2.8] | 2.5  [1.9–3.1] | 2.8  [2.2–3.4] | 3.0  [2.4–3.6] | 2.2  [1.6–2.8] |
| 35-44 | 0.4  [0.1–0.6] | 0.5  [0.2–0.8] | 0.4  [0.2–0.7] | 0.2  [0.0–0.4] | 0.3  [0.1–0.5] | 0.4  [0.2–0.7] | 1.5  [1.0–2.1] | 1.5  [1.0–1.9] | 2.1  [1.5–2.7] | 1.8  [1.3–2.3] | 1.6  [1.1–2.0] |
| 45-54 | 0.6  [0.3–0.8] | 0.5  [0.2–0.7] | 0.4  [0.2–0.7] | 0.3  [0.1–0.5] | 0.4  [0.2–0.6] | 0.3  [0.1–0.5] | 1.2  [0.8–1.7] | 1.3  [0.9–1.7] | 1.8  [1.3–2.3] | 1.5  [1.0–1.9] | 1.3  [0.8–1.8] |
| 55-64 | 0.4  [0.2–0.6] | 0.5  [0.2–0.8] | 0.5  [0.2–0.7] | 0.4  [0.1–0.6] | 0.4  [0.2–0.6] | 0.5  [0.2–0.8] | 0.7  [0.4–1.1] | 0.9  [0.6–1.3] | 1.5  [1.0–1.9] | 1.5  [1.1–2.0] | 1.0  [0.7–1.4] |
| ≥65 | 0.5  [0.3–0.7] | 0.4  [0.2–0.6] | 0.6  [0.4–0.8] | 0.4  [0.3–0.6] | 0.6  [0.4–0.8] | 0.4  [0.2–0.6] | 0.9  [0.6–1.1] | 1.3  [1.0–1.6] | 1.1  [0.8–1.4] | 0.9  [0.6–1.2] | 0.8  [0.6–1.0] |
