## Supplementary file 4 for "The changing face of nicotine use in England: age-specific annual trends, 2014-2024"

### Table S18. Proportion who have never regularly smoked among adults who vape, by age group and year

|  | **Never regularly smoked, % [95% confidence interval]** | | | | | | | | | | |
| --- | --- | --- | --- | --- | --- | --- | --- | --- | --- | --- | --- |
|  | **2014** | **2015** | **2016** | **2017** | **2018** | **2019** | **2020** | **2021** | **2022** | **2023** | **2024** |
| 18-24 | 4.3  [0.9–7.6] | 1.9  [0.0–4.2] | 2.6  [0.3–5.0] | 5.5  [1.4–9.5] | 7.2  [2.3–12.1] | 9.2  [4.0–14.4] | 15.1  [9.4–20.8] | 14.3  [9.3–19.3] | 19.1  [15.0–23.3] | 28.6  [24.3–33.0] | 34.3  [29.8–38.8] |
| 25-34 | 1.5  [0.0–3.2] | 3.9  [0.9–6.9] | 4.2  [1.5–6.9] | 8.4  [4.4–12.4] | 3.2  [0.6–5.9] | 9.1  [5.0–13.2] | 5.4  [2.4–8.4] | 4.5  [2.1–6.9] | 10.3  [7.3–13.3] | 11.1  [8.2–14.0] | 13.3  [10.3–16.2] |
| 35-44 | 4.5  [1.1–8.0] | 1.0  [0.0–2.2] | 5.4  [2.2–8.7] | 7.3  [3.3–11.4] | 7.2  [3.4–10.9] | 5.7  [2.3–9.0] | 2.7  [0.5–4.9] | 4.8  [1.9–7.8] | 5.9  [3.1–8.8] | 8.4  [5.4–11.5] | 9.6  [6.4–12.8] |
| 45-54 | 2.9  [0.5–5.2] | 5.8  [2.0–9.6] | 3.9  [1.2–6.6] | 5.6  [2.4–8.7] | 6.2  [2.8–9.6] | 7.0  [3.5–10.4] | 5.0  [2.1–8.0] | 3.7  [1.2–6.3] | 7.0  [3.7–10.4] | 5.6  [2.8–8.4] | 6.3  [3.3–9.3] |
| 55-64 | 1.6  [0.0–3.5] | 1.6  [0.0–3.3] | 4.8  [1.5–8.2] | 3.3  [0.7–5.9] | 5.1  [1.6–8.7] | 4.8  [1.7–7.9] | 6.5  [2.5–10.4] | 3.1  [0.4–5.9] | 1.6  [0.0–3.4] | 5.1  [2.2–7.9] | 4.5  [1.7–7.3] |
| ≥65 | 1.9  [0.0–4.1] | 6.2  [0.0–12.4] | 4.5  [0.9–8.2] | 3.8  [0.2–7.5] | 5.4  [1.3–9.5] | 7.9  [2.8–13.1] | 8.3  [3.7–13.0] | 4.6  [0.5–8.7] | 3.8  [0.0–7.7] | 2.1  [0.0–4.3] | 4.2  [0.5–7.9] |

### Table S19. Proportion who quit smoking ≥1 year ago among adults who vape, by age group and year

|  | **Quit smoking ≥1 year ago, % [95% confidence interval]** | | | | | | | | | | |
| --- | --- | --- | --- | --- | --- | --- | --- | --- | --- | --- | --- |
|  | **2014** | **2015** | **2016** | **2017** | **2018** | **2019** | **2020** | **2021** | **2022** | **2023** | **2024** |
| 18-24 | 5.1  [0.9–9.3] | 5.5  [1.5–9.5] | 7.0  [3.1–10.9] | 9.1  [4.3–13.9] | 7.2  [2.9–11.5] | 13.8  [7.1–20.5] | 12.9  [7.3–18.5] | 14.6  [9.6–19.7] | 13.1  [9.5–16.7] | 12.4  [9.0–15.9] | 19.3  [15.4–23.1] |
| 25-34 | 10.7  [6.0–15.3] | 14.9  [9.6–20.1] | 15.8  [10.9–20.7] | 23.7  [17.8–29.6] | 25.5  [19.6–31.4] | 25.7  [19.2–32.1] | 31.5  [25.3–37.6] | 26.4  [21.1–31.8] | 28.9  [24.3–33.4] | 31.0  [26.7–35.3] | 35.5  [31.3–39.7] |
| 35-44 | 9.6  [5.1–14.0] | 18.8  [12.9–24.7] | 24.4  [17.7–31.2] | 28.6  [21.9–35.2] | 27.4  [20.6–34.1] | 36.2  [28.3–44.0] | 47.4  [39.9–54.9] | 36.8  [29.7–43.9] | 42.9  [36.6–49.1] | 37.7  [32.6–42.9] | 44.8  [39.6–49.9] |
| 45-54 | 10.9  [6.2–15.5] | 14.7  [9.7–19.6] | 26.7  [20.5–32.9] | 29.9  [23.4–36.4] | 35.2  [28.3–42.2] | 35.7  [28.4–43.0] | 38.6  [31.5–45.8] | 42.5  [35.6–49.4] | 38.3  [31.9–44.7] | 48.9  [43.2–54.7] | 45.9  [39.8–52.0] |
| 55-64 | 15.7  [8.5–23.0] | 13.2  [7.5–18.9] | 25.2  [18.1–32.3] | 33.8  [26.2–41.3] | 30.1  [22.6–37.5] | 39.9  [32.1–47.6] | 45.0  [36.9–53.1] | 44.6  [37.2–52.1] | 41.9  [33.8–50.0] | 40.6  [34.4–46.8] | 54.2  [47.5–60.8] |
| ≥65 | 13.9  [7.0–20.8] | 19.1  [11.0–27.2] | 24.2  [16.3–32.1] | 32.8  [23.9–41.8] | 41.7  [32.8–50.6] | 37.1  [27.4–46.8] | 40.3  [30.7–49.9] | 46.3  [37.2–55.4] | 48.2  [37.9–58.4] | 52.1  [43.5–60.8] | 55.2  [47.1–63.3] |

### Table S20. Proportion who quit smoking <1 year ago among adults who vape, by age group and year

|  | **Quit smoking <1 year ago, % [95% confidence interval]** | | | | | | | | | | |
| --- | --- | --- | --- | --- | --- | --- | --- | --- | --- | --- | --- |
|  | **2014** | **2015** | **2016** | **2017** | **2018** | **2019** | **2020** | **2021** | **2022** | **2023** | **2024** |
| 18-24 | 6.2  [2.1–10.3] | 10.2  [4.8–15.7] | 9.9  [5.4–14.4] | 10.5  [5.1–15.9] | 12.6  [6.1–19.1] | 5.5  [1.7–9.3] | 9.9  [5.0–14.8] | 11.9  [7.4–16.4] | 13.2  [9.6–16.9] | 9.8  [6.8–12.9] | 8.5  [5.8–11.3] |
| 25-34 | 10.6  [5.8–15.4] | 10.2  [5.7–14.7] | 14.4  [8.8–20.0] | 10.8  [6.1–15.6] | 9.2  [5.4–13.0] | 6.8  [2.6–11.0] | 10.6  [6.8–14.4] | 12.5  [8.5–16.4] | 10.5  [7.6–13.3] | 11.4  [8.6–14.3] | 11.7  [8.8–14.7] |
| 35-44 | 18.9  [12.7–25.1] | 13.1  [7.8–18.3] | 11.4  [6.8–16.0] | 7.0  [3.2–10.7] | 8.6  [4.2–13.0] | 4.7  [1.0–8.5] | 8.9  [4.8–13.1] | 9.4  [5.0–13.8] | 10.8  [6.9–14.8] | 11.3  [7.9–14.8] | 10.0  [6.9–13.0] |
| 45-54 | 10.9  [6.0–15.9] | 13.6  [8.5–18.8] | 10.4  [6.1–14.7] | 11.1  [6.5–15.7] | 6.4  [3.0–9.8] | 7.2  [3.3–11.1] | 7.3  [3.3–11.2] | 8.0  [4.1–11.8] | 7.4  [4.0–10.9] | 6.0  [3.5–8.6] | 6.4  [3.6–9.2] |
| 55-64 | 9.0  [3.8–14.3] | 8.7  [4.0–13.4] | 9.6  [4.7–14.4] | 8.2  [4.0–12.4] | 4.9  [1.5–8.4] | 5.5  [1.5–9.6] | 1.7  [0.0–3.4] | 7.8  [3.8–11.8] | 6.5  [2.3–10.6] | 8.1  [4.4–11.9] | 6.8  [3.6–10.1] |
| ≥65 | 10.5  [4.9–16.2] | 8.1  [1.8–14.3] | 11.3  [5.2–17.5] | 8.1  [3.1–13.1] | 1.2  [0.0–2.8] | 3.9  [0.4–7.3] | 3.6  [0.0–7.1] | 4.0  [0.0–8.0] | 10.6  [4.7–16.5] | 6.6  [2.1–11.1] | 5.1  [1.4–8.8] |

### Table S21. Proportion who currently smoke among adults who vape, by age group and year

|  | **Currently smoke, % [95% confidence interval]** | | | | | | | | | | |
| --- | --- | --- | --- | --- | --- | --- | --- | --- | --- | --- | --- |
|  | **2014** | **2015** | **2016** | **2017** | **2018** | **2019** | **2020** | **2021** | **2022** | **2023** | **2024** |
| 18-24 | 84.4  [78.1–90.8] | 82.4  [75.7–89.2] | 80.5  [74.5–86.5] | 74.9  [67.3–82.5] | 72.9  [64.7–81.2] | 71.6  [63.4–79.7] | 62.1  [54.0–70.2] | 59.2  [52.2–66.1] | 54.5  [49.3–59.8] | 49.1  [44.3–54.0] | 37.9  [33.3–42.5] |
| 25-34 | 77.3  [71.0–83.6] | 71.1  [64.4–77.8] | 65.5  [58.6–72.4] | 57.1  [50.0–64.1] | 62.1  [55.5–68.6] | 58.4  [51.2–65.6] | 52.6  [45.9–59.2] | 56.6  [50.6–62.6] | 50.4  [45.4–55.4] | 46.4  [41.7–51.1] | 39.4  [35.0–43.9] |
| 35-44 | 67.0  [59.6–74.4] | 67.2  [60.0–74.3] | 58.7  [51.3–66.1] | 57.2  [49.9–64.4] | 56.9  [49.4–64.3] | 53.4  [45.4–61.5] | 41.0  [33.5–48.4] | 48.9  [41.6–56.3] | 40.4  [34.2–46.5] | 42.5  [37.2–47.7] | 35.7  [30.6–40.7] |
| 45-54 | 75.3  [68.7–81.9] | 65.9  [59.0–72.9] | 59.0  [52.1–65.8] | 53.5  [46.5–60.6] | 52.2  [45.1–59.3] | 50.1  [42.4–57.8] | 49.1  [41.6–56.6] | 45.8  [38.8–52.8] | 47.2  [40.6–53.9] | 39.4  [33.8–45.0] | 41.4  [35.4–47.5] |
| 55-64 | 73.6  [65.1–82.1] | 76.5  [69.4–83.5] | 60.4  [52.5–68.3] | 54.7  [46.8–62.6] | 59.9  [51.9–67.8] | 49.8  [42.0–57.7] | 46.8  [38.7–54.9] | 44.4  [36.9–52.0] | 50.0  [41.7–58.4] | 46.2  [39.8–52.5] | 34.5  [28.1–40.9] |
| ≥65 | 73.7  [65.1–82.2] | 66.6  [56.2–77.0] | 60.0  [50.8–69.2] | 55.3  [45.8–64.7] | 51.8  [42.7–60.8] | 51.1  [41.1–61.0] | 47.8  [38.1–57.5] | 45.1  [36.0–54.2] | 37.4  [27.6–47.2] | 39.1  [30.6–47.6] | 35.5  [27.7–43.4] |
